# Arteriovenous Sampling for Organ-Specific Metabolic Insights in CKM Syndrome

**DOI:** 10.1101/2025.06.24.25330247

**Authors:** Dana Hicks, Anu Joyson, Natalie C. Ward, Bu B. Yeap, Girish Dwivedi, Julien Wist, Markus Schlaich, Nicola Gray

## Abstract

Cardiovascular-kidney-metabolic (CKM) syndrome recognises connections among obesity, diabetes, chronic kidney disease, and cardiovascular disease, describing the close metabolic links and shared risk factors including dyslipidemia, hypertension, and metabolic dysfunction. While the concentrations of circulating metabolites have been frequently measured in these varied pathophysiological conditions, the flux of metabolites between and across organs is not easily captured. Arteriovenous sampling provides a transformative approach to metabolite gradient measurements offering new insights into organ-specific regulation of lipid, lipoprotein, and small molecule metabolism.

Here, arteriovenous sampling was used to determine organ-specific metabolic differences between the heart and kidney compared with peripheral venous differences. Liquid chromatography-tandem mass spectrometry (LC-MS/MS) and nuclear magnetic resonance (NMR) analysis was performed to assess lipids, lipoproteins and small molecules in peripheral venous, coronary sinus (CS), renal vein and radial artery samples collected from 18 participants with stage 1-2 CKM.

Paired arterial-CS and arterial-renal vein gradients revealed significant differences in lipid and lipoprotein metabolism. The heart exhibited net uptake of long-chain fatty acids (e.g., FA(18:0), FA(20:3)) and very-low-density lipoprotein (VLDL) subfractions (e.g., VLFC, VLAB), along with a net release of lysophosphatidylethanolamine (LPE(18:2)). In contrast, the kidney showed net uptake of diacylglycerols (DG), fatty acids (e.g., FA(14:0), FA(16:1)), and lysophospholipids (e.g., LPE(20:4)). Significant differences in glutamine, citric acid, lactic acid, and pyruvic acid gradients between the heart and kidneys were noted, with each analyte showing a net uptake by its respective organ.

Our findings highlight the comprehensive changes in organ-specific metabolites and lipids that occur between the heart and kidneys. These data indicate the value of arteriovenous sampling in understanding the metabolic differences between the heart and kidneys, and its potential to explore bidirectional metabolic crosstalk between these organs.

## Background

The cardiovascular and renal systems are closely linked through metabolic processes, with diseases often arising from shared pathophysiological mechanisms and risk factors such as obesity, dysglycemia, dyslipidemia, and blood pressure [1]. Cardiovascular-kidney-metabolic (CKM) syndrome, as defined by the American Heart Association, encompasses interrelated health conditions including obesity, diabetes, chronic kidney disease, and cardiovascular disease [2]. These conditions share common aetiologies, including insulin resistance, altered blood pressure regulation and dysregulated lipid metabolism [2].

Obesity has been associated with insulin resistance, hypertension, dyslipidemia, and chronic inflammation, all of which are central to CKM syndrome [2,3]. Excess abdominal adiposity plays a critical role in the pathophysiology of CKM syndrome [2,3]. Abdominal obesity is commonly assessed using body mass index (BMI) and a waist circumference (WC) ≥102 cm in men and ≥88 cm in women, with lower thresholds for Asian populations (≥90 cm in men and ≥80 cm in women) [2,4]. The expansion of white adipose tissue recruits inflammatory cells and remodels vasculature, leading to white adipose tissue dysfunction [5]. This dysfunction suppresses adipokine secretion, alters lipoprotein functionality, and impairs lipid transport and storage, which together exacerbate inflammation [5,6]. Consequently, excess lipids enter the bloodstream, leading to lipid toxicity and metabolic strain in the heart and kidneys due to altered nutrient utilisation and impaired mitochondrial function [7]. This lipotoxicity impairs heart and kidney function, accelerating CKM syndrome progression [2,5]. However, the mechanistic pathways underlying the bidirectional relationship between organ-specific dysfunction and the reciprocal impact each organ has on the other remain poorly understood [8]. Bridging these gaps is essential for tailoring treatments to disease progression and addressing the clinical heterogeneity of CKM patients [8].

Comprehensive metabolic phenotyping enables the characterisation of lipids, lipoproteins, and small molecules, providing insights into energy production, metabolic flexibility, mitochondrial function, and lipid metabolism in CKM syndrome [8,9]. Similarly, arteriovenous sampling across the organ of interest serves as an investigative tool to uncover organ-specific metabolic exchange, offering a deeper understanding of cardiovascular-renal metabolic interactions [10]. Arteriovenous sampling involves simultaneous blood collection from both venous and arterial circulations around individual organs, allowing for the direct comparison of metabolite concentrations in venous and arterial blood [10]. This technique can assess metabolite uptake or release by organs or tissues, with the arteriovenous gradient (AVG) calculated using the log ratio of metabolite concentrations [10]. An AVG > 0 indicates metabolite release, while an AVG < 0 suggests metabolite uptake [10].

While peripheral sampling provides a broad view of metabolism, it is influenced by factors such as diet, hormonal regulation, and metabolic adaptations, which limit the ability to examine organ-specific metabolic processes [9]. To address this, arteriovenous sampling of the coronary sinus (CS) and renal veins enables the individual assessment of cardiac and renal metabolism while also allowing for the integrated analysis of their bidirectional metabolic crosstalk. This approach enhances understanding of organ-specific metabolic processes in CKM syndrome, particularly in the context of cardiovascular and renal metabolism [10].

This study aimed to use comprehensive metabolite phenotyping using LC-MS/MS and NMR applied to arteriovenous sampling to identify organ-specific metabolic differences between the heart and kidneys, and explore differences compared with peripheral venous sampling. The benefits of arteriovenous sampling were investigated on samples collected from an ongoing trial of sodium glucose cotransporter-2 (SGLT2) inhibition in participants with stage 1-2 CKM.

## Material and methods

### Study Design and Participants

Samples were analysed from 18 participants of a randomised, double-blinded, placebo-controlled, crossover study of SGLT2 inhibition and sympathetic nervous system activity (ClinicalTrials.gov ID NCT03912909). All met the following criteria: metabolic syndrome, defined as having obesity (BMI ≥30 kg/m²) plus any two of the following four factors: triglycerides ≥1.7 mmol/L, high-density lipoprotein (HDL) <1.03 mmol/L (men) or <1.29 mmol/L (women), systolic BP ≥130 or diastolic BP ≥85 mm Hg and fasting glucose ≥5.6 mmol/L. Additionally, all participants had a waist circumference (WC) ≥102 cm (men) or ≥88 cm (women), further aligning with stage 1-2 CKM syndrome [2]. Baseline physical and biochemical characteristics were similar for eight female and 10 male participants (Table S1).

Participants received empagliflozin or placebo daily for 4 weeks, followed by a 4-week washout, then reversed treatments for another 4 weeks. The Royal Perth Hospital Pharmacy handled randomisation, blinding, and medication distribution. At the end of each 4-week treatment phase, coronary sinus (CS) and right renal vein (RRV) samples were obtained with simultaneous sampling from the radial artery (Figure 1). Central venous catheter placement was performed via an 8.5F percutaneous introducing sheath into the femoral or antecubital vein. Venous blood samples were collected from the CS, followed by the RRV 8.3 minutes (mean) later. Each venous sample was paired with a simultaneous arterial sample from the radial artery. The arterial sample drawn from the radial artery is designated as A1 when paired with the CS and A2 with the RRV. Peripheral blood samples were obtained via a peripheral vein (PV) before any catheter procedure. Samples were transferred to GSH (glutathione)-containing tubes, stored on ice, centrifuged at 358 g for 40 minutes at 4°C, and stored at –80°C until analysis.

**Figure 1.**
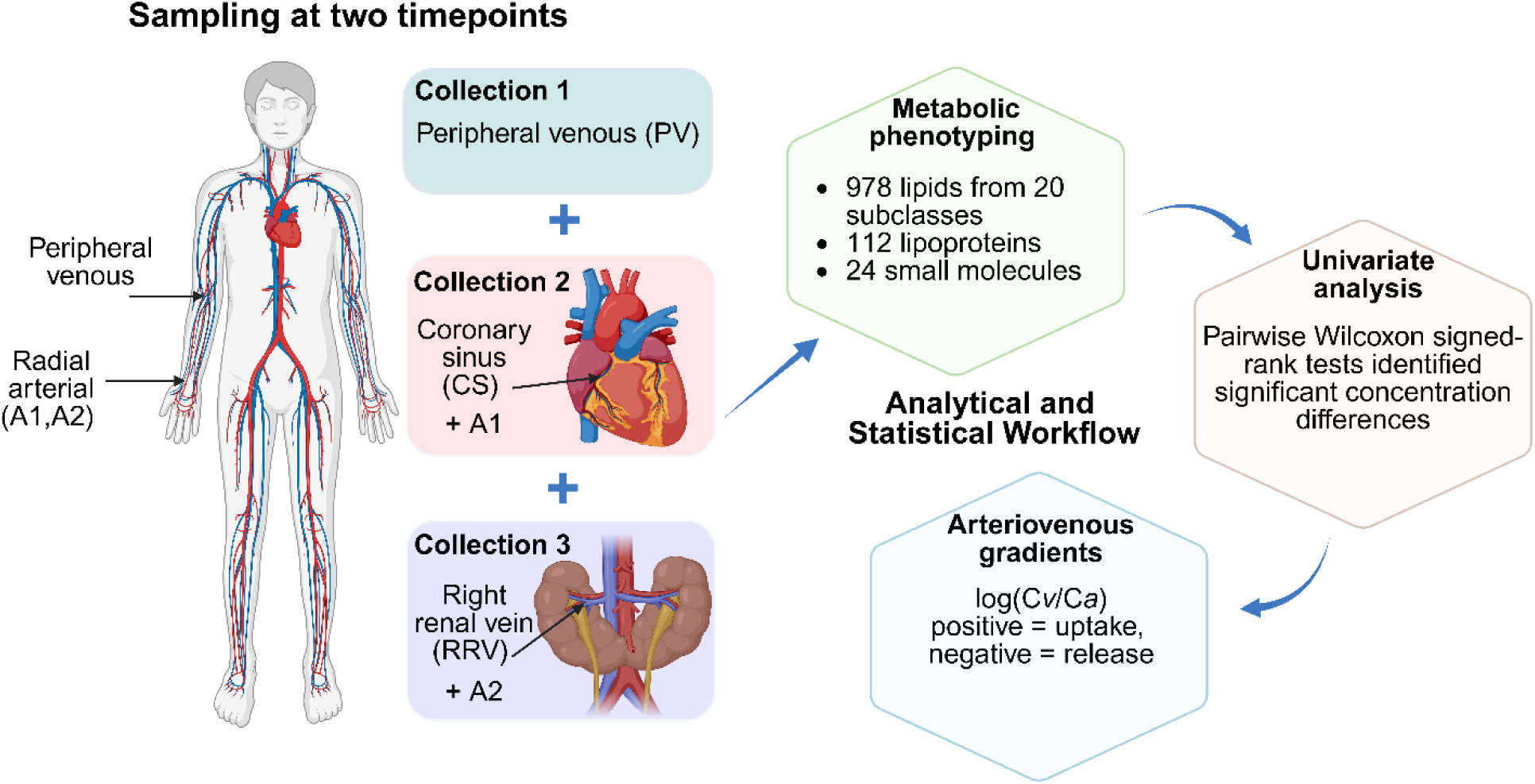
Overview of Sample Collection, Analytical, and Statistical Workflow. Samples were collected at two treatment time points, including peripheral venous (PV) samples and organ-specific arteriovenous sampling. For the heart, venous blood was collected from the coronary sinus (CS) with simultaneous radial artery sampling (A1). For the kidney, venous blood was collected from the right renal vein (RRV) with simultaneous radial artery sampling (A2). Metabolic phenotyping included targeted lipid profiling by LC-MS/MS and analysis of lipoproteins and small molecules by NMR spectroscopy. Univariate analysis identified significant concentration differences between RRV and PV (RRV∼PV), CS and PV (CS∼PV), CS and RRV (CS∼RRV), A1 and A2 (A1∼A2), A2 and RRV (A2∼RRV) and A1 and CS (A1∼CS). Arteriovenous gradients were calculated as the log-transformed ratio of venous to arterial concentrations [log(Cv/Ca)], where positive values indicate net organ uptake and negative values indicate net release. For the heart, gradients were derived using mean concentrations from CS and A1; for the kidney, RRV and A2.

### Ethical Approval

Ethical approval was granted by the Royal Perth Hospital Human Research Ethics Committee (Ref: 16-157) and Murdoch University Human Research Ethics Committee (Project: 2023/165). All participants provided written informed consent.

### Lipidomic Analysis using LC-MS/MS

Plasma samples were analysed using a previously described protocol for detecting 1163 lipid species [11]. Briefly, 20 μL of plasma was vortex-mixed with 180 μL of isopropanol containing stable isotope-labelled internal standards (ISTD) including UltimateSPLASH^TM^ ONE, SphingoSPLASH^TM^ I, 17:1 Lyso PS, 18:1-d7 MG (Sigma-Aldrich, North Ryde, NSW, Australia), linoleic acid-d_11_, arachidonic acid-d_5_, palmitic acid-d_5_, stearic acid-d_4_ (Sapphire Bioscience, Redfern, NSW, Australia). Samples were centrifuged at 14 000*g* for 15 min, and the supernatant was transferred to a 96-well plate. Pooled quality control (PQC) samples were created by combining an aliquot from each study sample. A commercial plasma long-term reference (LTR) sample (BioIVT, Westbury, NY, USA) was also prepared. Both PQC and LTR samples were processed alongside the study samples.

The reversed-phase LC-MS/MS was performed using an Acquity Premier BEH C18 1.7 µm, 2.1 x100 mm column (Waters Corporation, MA, USA) with an ExionLC^TM^ coupled to a QTRAP 6500+ mass spectrometer (SCIEX, Concord, CA). The PQC and LTR were analysed throughout the entire analytical run as a quality control strategy to assess analytical stability [12]. A full description is provided in Supplementary Box 1.

### Lipidomic Data Pre-processing

An in-house custom R pipeline (v4.4.1, R Foundation, Vienna, Austria) within R Studio (v1.4.1, R Studio, Boston, MA, USA) was utilised to pre-process the LC-MS/MS data obtained from SCIEX OS 3.0 (SCIEX, MA, USA). This pre-processing included peak picking using SkylineMS 21.1 (MacCoss, Seattle, WA, USA), evaluating data quality, calculating relative concentrations by determining the area under the peak relative to the ISTD used and additional quality control steps as previously described [11].

### *1H* NMR Spectroscopic Data Acquisition and Processing

Plasma samples (300 μL) were prepared by mixing with 300 μL of phosphate buffer (75 mM Na_2_HPO_4_, 2 mM NaN_3_, 4.6 mM sodium trimethylsilyl propionate-[2,2,3,3-^2^H_4_] (TSP) in H_2_O/D_2_O 4:1, pH 7.4 ± 0.1) were transferred to 5 mm NMR SampleJet tubes and sealed with POM balls. Samples were manually shaken to remove bubbles and stored inside the SampleJet automatic sample changer for analysis.

NMR spectroscopic analyses were conducted using a 600 MHz Bruker Avance III HD spectrometer (Bruker BioSpin, Billerica, Massachusetts, USA) equipped with a 5 mm broadband inverse (BBI) probe and a SampleJet robotic cooling system Bruker BioSpin, Billerica, Massachusetts, USA) set at 5 °C. Before analysis, a full quantitative calibration was performed following a previously described protocol [13]. A single one-dimensional (1D) NMR experiment was performed for each sample using Bruker’s In Vitro Diagnostics research (IVDr) methods. This method utilised a 90° pulse and solvent pre-saturation, acquiring 32 scans with 98,000 data points across a spectral width of 30 parts per million (ppm). The total experiment duration was 4 minutes and 3 seconds [13]. Lipoprotein analysis was conducted using the Bruker IVDr Lipoprotein Subclass Analysis (B.I.-LISA^TM^) method to quantify 112 lipoprotein parameters (Table S2) [14], with the full description provided in Box S2. Additionally, concentrations of 24 small molecules (molecular weight < 1,500 Da) were quantified using the Bruker IVDr Quant-PS^TM^ protocol (Table S3) [14].

### Data Analysis

Data analysis was performed using R (v4.4.1) with plots generated through *ComplexUpset* (v1.3.3), and *ggplot2* (v3.5.1). Data from the same participants at two time points were combined to avoid treatment influence. Univariate analysis was performed on the concentration data of venous samples to identify significant differences between the sampling sites. The Friedman test, which accounts for paired participant samples across time points, was used to reduce treatment-related variability. Significant lipids identified by the Friedman test were further analysed using post-hoc pairwise Wilcoxon signed-rank tests (*stats* package), with p-values adjusted using the Benjamini–Hochberg method (p < 0.05).

Pairwise comparisons between sampling sites were used to explore organ-specific and systemic metabolic differences. These included comparisons between renal vein (RRV), coronary sinus (CS), peripheral vein (PV) and arterial samples (A1, A2). Arteriovenous gradients (AVG) were calculated as the log-transformed ratio of venous to arterial concentrations: log(Cv/Ca) [10,15]. For the heart, CS samples were paired with A1; for the kidney, RRV was paired with A2. A positive AVG indicates net uptake, while negative AVG indicates net release [10,15]. Mean AVG and 95% confidence intervals (CI) were used to assess metabolite exchange. CI = Mean ± (*t* × SE), where *t* is the critical value and SE is the standard error. Organ-specific AVG differences identified lipoprotein, lipid and small molecule utilisation or release.

## Results

Comprehensive LC-MS/MS and NMR metabolic phenotyping was performed on arterial (A1,A2), peripheral venous (PV), coronary sinus (CS), and right renal vein (RRV) samples collected from 18 participants at two time points (Figure 1).

Univariate comparisons across sampling sites revealed significant concentration differences reflecting metabolic activity in the heart, kidney and systemic circulation. Specifically, RRV∼PV and CS∼PV comparisons highlighted kidney– and heart-associated venous signatures relative to systemic circulation. A1∼A2 comparisons assessed temporal changes in arterial metabolite levels. Comparisons between A2∼RRV and A1∼CS examined arterial-to-venous concentration differences across the kidney and heart, respectively, while CS∼RRV enabled direct comparison of venous effluent from both organs.

To quantify organ-specific metabolite exchange, AVGs were calculated for the heart and kidney using paired arterial and venous samples. These gradients provide direct insight into net uptake or release of metabolites by each organ, offering a more physiologically meaningful measure of organ-specific metabolism than concentration comparisons alone.

### Lipidomic Univariate Analysis

Across all sampling sites, 504 lipids showed significant concentration differences between PV, CS, and RRV samples, while 285 significant lipids were identified in the arterial (A1∼A2) comparisons (Table 1). The direction of these lipid concentration differences, indicating increases or decreases, is outlined in Table S4. Interestingly, we observed distinct patterns in lipid species concentrations across sampling sites. When comparing RRV and CS samples to PV (RRV∼PV and (CS∼PV), multiple species within the diacylglycerol (DG), fatty acyl (FA), lysophosphatidylcholine (LPC), LPE, lysophosphatidylglycerol (LPG), lysophosphatidylinositol (LPI), lysophosphatidylserine (LPS), and monoacylglycerol (MG) subclasses showed consistently lower concentrations in PV. In contrast, species within the phosphatidylethanolamine (PE), phosphatidylglycerol (PG), phosphatidylserine (PS), and TG subclasses were elevated in PV relative to RRV and CS (Table S4).

**Table 1.** Number of Significant Lipids by Class across RRV∼PV, CS∼PV, CS∼RRV, A1∼A2, A2∼RRV, and A1∼CS comparisons

| Subclass | RRV~PV | CS~PV | CS~RRV | A1~A2 | A2~RRV | A1~CS |
| --- | --- | --- | --- | --- | --- | --- |
| CE | 4 | 4 | 0 | 0 | 0 | 0 |
| DG | 15 | 16 | 2 | 1 | 1 | 0 |
| FA | 20 | 19 | 17 | 16 | 5 | 8 |
| LPC | 10 | 10 | 4 | 10 | 0 | 0 |
| LPE | 14 | 14 | 5 | 12 | 2 | 1 |
| LPG | 6 | 5 | 4 | 5 | 0 | 0 |
| LPI | 7 | 7 | 3 | 5 | 1 | 0 |
| LPS | 7 | 5 | 5 | 4 | 0 | 0 |
| MG | 13 | 13 | 1 | 12 | 0 | 0 |
| PE | 9 | 9 | 0 | 1 | 0 | 0 |
| PG | 3 | 3 | 0 | 0 | 0 | 0 |
| PS | 4 | 4 | 1 | 1 | 0 | 0 |
| TG | 386 | 380 | 234 | 218 | 0 | 0 |
| **Total** | **498** | **489** | **276** | **285** | **9** | **9** |
| Note: Lipid subclass key- cholesterol ester (CE), diacylglycerol (DG), fatty acyl (FA), lysophosphatidylcholine (LPC), lysophosphatidylethanolamine (LPE), lysophosphatidylglycerol (LPG), lysophosphatidylinositol (LPI), lysophosphatidylserine (LPS), monoacylglycerol (MG), phosphatidylethanolamine (PE), phosphatidylglycerol (PG), phosphatidylserine (PS), triacylglycerols (TG) |  |  |  |  |  |  |

Distinct patterns also emerged when comparing arterial samples A1 (paired with CS) and A2 (paired with RRV), primarily from the glycerolipids, glycerophospholipid, and FA subclasses (Table S4). Notably, the following lipid species showed increased concentrations in the arterial samples, including DG(18:2_20:4), FA(14:1), FA(16:2), LPC(18:3), LPC(20:2), LPC(20:3), LPC(20:4), LPC(22:5), LPC(22:6), LPE(18:1), LPE(18:2), LPE(18:3), LPE(20:3), LPE(20:4), LPE(22:6), LPG(18:1), LPG(18:2), LPI(16:1), LPI(18:2), and LPS(16:0). All MG species showed increased concentrations in the A1∼A2 comparison except for MG(22:3), which did not exhibit a distinct trend. In contrast, the following lipid species showed decreased concentrations exclusively in the arterial samples, including TG(48:3_FA14:0), TG(48:3_FA16:0), TG(54:5_FA16:1), TG(56:4_FA22:4), TG(56:6_FA20:5), and TG(56:7_FA18:3). In contrast, other lipids exhibited coordinated changes in the A1∼A2 and CS∼RRV samples from all the lipid subclasses outlined in Table 1 except for cholesterol ester (CE), DG, PE and PG (Table S4).

Both the A2∼RRV and A1∼CS arteriovenous comparisons identified nine significant lipids each (Table 1). Distinct lipid profiles were observed in these comparisons, reflecting differences in lipid metabolism within the kidney and the heart, respectively. In the A2∼RRV comparison, significant lipids such as DG(14:0_20:4), FA(14:0), FA(14:1), FA(16:1), FA(16:2), FA(18:3), LPE(22:6), and LPI(16:1) showed lower concentrations in RRV compared to A2 (Table S4). In contrast, the A1∼CS comparison revealed reduced concentrations of FA(18:0), FA(18:1), FA(20:0), FA(20:1), FA(20:2), FA(20:3), FA(22:4), and FA(22:5) in CS, while LPE(18:2) exhibited higher concentrations in CS than A1 (Table S4).

An UpSet plot visualised the intersections of significant lipids that differed between the following groups: RRV vs. PV, CS vs. PV, A1 vs. A2, CS vs. RRV, A2 vs. RRV, and A1 vs. CS (Figures 2A and 2B). The purpose of these comparisons is to identify how lipid profiles vary across these sampling sites. The horizontal bars (set size) represent the total number of significant lipids within each comparison, while the vertical bars (intersections) show the number of lipids shared across the comparisons. Dots connected by lines highlight overlapping lipids between these comparisons, while isolated dots indicate lipids unique to a specific comparison. This visualisation allows for the identification of both unique and shared lipid features, providing insights into the metabolic differences between the groups.

**Figure 2.**
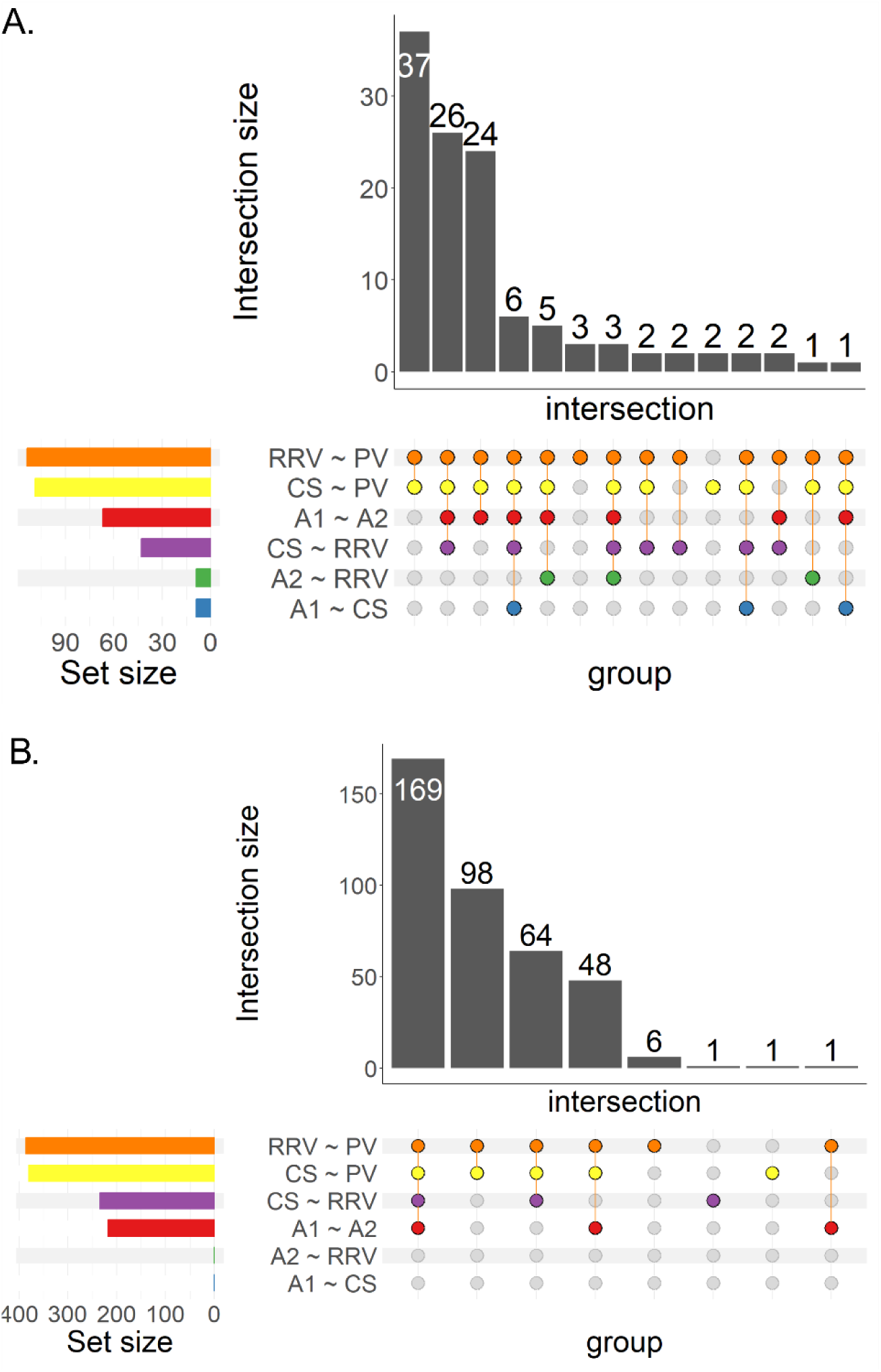
UpSet plot showing significant lipid distribution across RRV∼PV, CS∼PV, CS∼RRV, A1∼A2, A2∼RRV, and A1∼CS comparisons. A) DG, FA, LPC, LPE, LPG, LPI, LPS, MG, PE, and PS subclasses (Table S4), B) TG subclass (Table S4).

### Lipoprotein Univariate Analysis

Significant lipoproteins were identified through the pairwise Wilcoxon signed-rank tests across the RRV∼PV, CS∼PV, CS∼RRV, A1∼A2, A2∼RRV, and A1∼CS comparisons. Similar trends in lipoprotein concentration variations were observed when comparing the RRV and CS samples to PV, with both sampling sites predominantly showing coordinated increases or decreases. This was the only consistent pattern observed across the sampling site comparisons.

In the RRV∼PV comparison, 83 metabolites showed statistically significant concentration differences (adjusted *p*-value < 0.05), while the CS∼PV comparison revealed 73 such differences. Notably, LDL-free cholesterol (L5FC) and VLDL4-cholesterol (V4CH) were uniquely altered in the CS∼PV comparison. The direction of these lipoprotein concentration differences, indicating increases or decreases, is outlined in Table S5. Comparisons between paired CS and RRV samples identified 28 non-unique significant lipoprotein concentration differences. Fifty significant lipoproteins were found to differ between A1 and A2, including unique variations in HDL-cholesterol (HDCH), HDL-apolipoprotein-A1 (HDA1), lipoprotein subfractions HDL1-free cholesterol (H1FC), HDL3-apolipoprotein-A2 (H3A2), low-density lipoprotein (LDL) free cholesterol (L1FC), VLDL5-cholesterol (V5CH), and VLDL5-phospholipids (V5PL) (Table S5). The A2∼RRV comparison identified three significant differences, while the A1∼CS comparison revealed 18 significant differences, none of which were unique (Table S5). Figure 3 presents an UpSet plot illustrating the intersections of significant lipoproteins across these comparisons.

**Figure 3.**
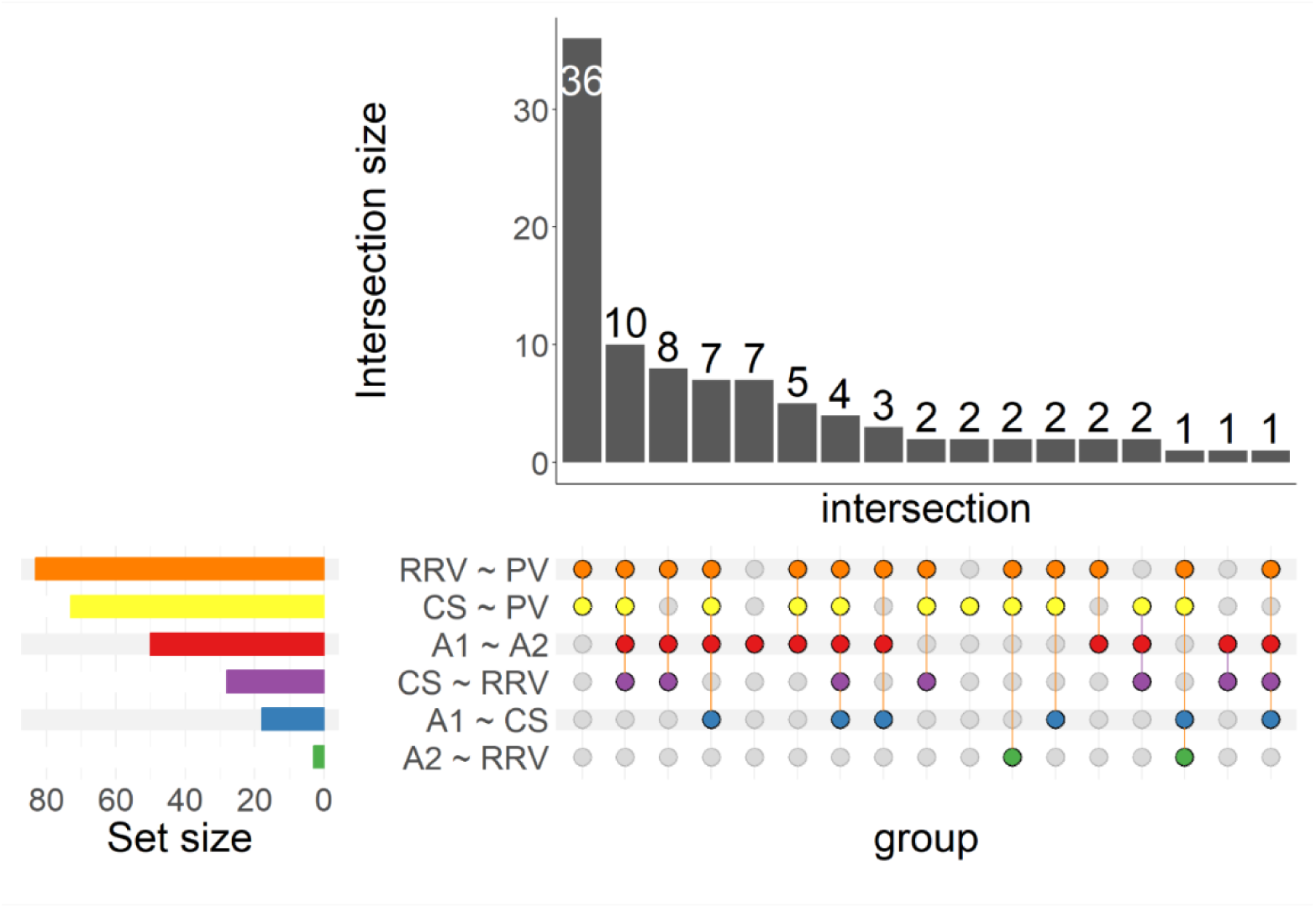
UpSet plot showing significant lipoproteins across RRV∼PV, CS∼PV, CS∼RRV, A1∼A2, A2∼RRV, and A1∼CS comparisons (Table S5).

### Small Molecule Univariate Analysis

In the analysis of small molecules, both overall and unique significant differences were identified (Table S6). Notably, the only consistent trend across sampling sites mirrored the lipid and lipoprotein results, where RRV and CS showed similar concentration changes when compared to PV (Table S6). Creatine was unique to the RRV∼PV comparison, while methionine was unique to the CS∼RRV comparison (Table S6). For the A2∼RRV comparison, glutamine, citric acid, and creatinine were identified as unique. Glutamic acid was unique to the A1∼CS comparison, and only citric acid differed significantly between A1 and A2 (Table S6). Figure 4 illustrates all intersections.

**Figure 4.**
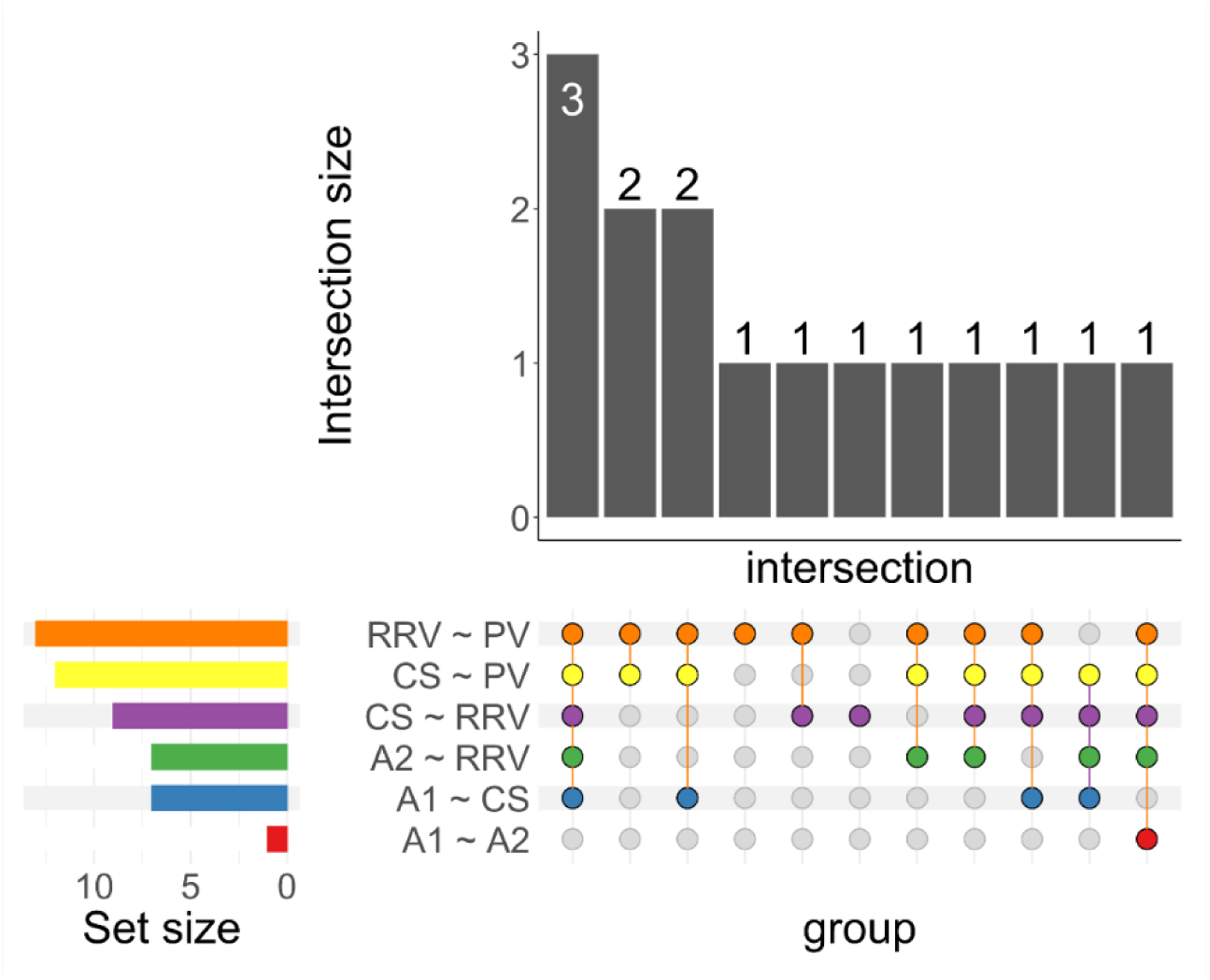
UpSet plot showing significant small molecules across RRV∼PV, CS∼PV, CS∼RRV, A1∼A2, A2∼RRV, and A1∼CS comparisons (Table S6).

### Arteriovenous Gradients

Analysis of AV gradients revealed significant differences in lipid, lipoprotein, and small molecule uptake or release by the heart and kidney (Table 2). Mean AV gradients and 95% confidence intervals for significant analytes in the A1∼CS and A2∼RRV comparisons illustrate net organ uptake or release. Significant uptake of long-chain FAs, including FA(18:0), FA(18:1), FA(20:0), FA(20:1), FA(20:2), FA(20:3), FA(22:4), and FA(22:5), was evident in the heart, alongside the release of LPE(18:2) (Table 2). In contrast, the kidney exhibited uptake of DG(16:0_20:4), FA(14:1), FA(16:1), FA(16:2), FA(18:3), and lysophospholipids LPE(20:4), LPE(22:6), and LPI(16:1) (Table 2).

**Table 2.** Arteriovenous gradients for each significant lipid, lipoprotein and small metabolite for A1∼CS and A2∼RRV representing net organ uptake (< 0) or release (> 0) by the heart or kidney.

| Analyte | Organ | Mean AV gradient | 95% CI |
| --- | --- | --- | --- |
| FA(18:0) | Heart | -0.12 | (-0.23, -0.02) |
| FA(18:1) | Heart | -0.15 | (-0.25, -0.03) |
| FA(20:0) | Heart | -0.11 | (-0.19, -0.03) |
| FA(20:1) | Heart | -0.35 | (-0.46, -0.23) |
| FA(20:2) | Heart | -0.22 | (-0.33, -0.12) |
| FA(20:3) | Heart | -0.13 | (-0.24, -0.02) |
| FA(22:4) | Heart | -0.22 | (-0.32, -0.12) |
| FA(22:5) | Heart | -0.12 | (-0.23, -0.01) |
| LPE(18:2) | Heart | 0.09 | (0.01, 0.16) |
| DG(16:0_20:4) | Kidney | -0.07 | (-0.15, 0.02) |
| FA(14:1) | Kidney | -0.16 | (-0.28, -0.04) |
| FA(16:1) | Kidney | -0.12 | (-0.21, -0.03) |
| FA(16:2) | Kidney | -0.19 | (-0.32, -0.07) |
| FA(18:3) | Kidney | -0.09 | (-0.17, -0.01) |
| LPE(20:4) | Kidney | -0.12 | (-0.22, -0.03) |
| LPE(22:6) | Kidney | -0.10 | (-0.18, -0.01) |
| LPI(16:1) | Kidney | -0.13 | (-0.23, -0.02) |
| TPTG | Heart | -0.04 | (-0.08, 0.003) |
| VLPN | Heart | -0.03 | (-0.06, 0.005) |
| VLFC | Heart | -0.04 | (-0.08, 0.003) |
| VLAB | Heart | -0.03 | (-0.06, 0.005) |
| V1TG | Heart | -0.10 | (-0.17, -0.03) |
| V1CH | Heart | -0.08 | (-0.13, -0.03) |
| V1PL | Heart | -0.07 | (-0.14, -0.009) |
| Glutamic acid | Heart | -0.37 | (-0.59, -0.16) |
| Lactic acid | Heart | -0.23 | (-0.35, -0.12) |
| Acetoacetic acid | Heart | -1.24 | (-1.91, -0.58) |
| Pyruvic acid | Heart | -0.53 | (-0.77, -0.29) |
| Glutamine | Kidney | -0.12 | (-0.19, -0.05) |
| Citric acid | Kidney | -0.62 | (-0.75, -0.49) |
| Lactic acid | Kidney | -0.08 | (-0.16, -0.006) |
| Acetoacetic acid | Kidney | 0.43 | (0.06, 0.79) |
| Pyruvic acid | Kidney | -0.10 | (-0.19, -0.002) |

Lipoprotein analysis showed significant heart uptake of total triglycerides (TPTG) and VLDL subfractions VLDL particle number (VLPN), VLDL-Free cholesterol (VLFC), VLDL-Apolipoprotein B100 (VLAB), VLDL1-Triglycerides (V1TG), VLDL1-Cholesterol (V1CH), and VLDL1-Phospholipids (V1PL), while no significant lipoproteins were associated with the kidney (Table 2). Small molecule analysis indicated the uptake of glutamic acid, lactic acid, acetoacetic acid, and pyruvic acid by the heart. The kidney, in contrast, displayed uptake of glutamine, citric acid, lactic acid, and pyruvic acid while releasing acetoacetic acid (Table 2). These observations highlight organ-specific metabolic activity in the heart and kidney.

## Discussion

This study evaluates the effectiveness of arteriovenous sampling for investigating lipid, lipoprotein and small molecule metabolism in the heart and kidney while recognising its limitations for clinical practice. The venous comparisons (RRV∼PV, CS∼PV, CS∼RRV) revealed significant differences in lipid, lipoprotein, and small molecule concentrations, highlighting systemic metabolic variations. In contrast, A1∼CS and A2∼RRV comparisons provide more insight into the metabolism of the heart and kidney by capturing differences between arterial input and venous output. Notably, TGs were significantly increased in RRV∼PV and CS∼PV comparisons but decreased in CS∼RRV comparisons. Despite these differences, TGs did not contribute to the AVGs, which are calculated using the log(Cv/Ca) ratio to assess net uptake or release by organs. The lack of significant TG changes in the arterial-to-venous comparisons (A1∼CS and A2∼CS) indicates that the observed venous differences reflect systemic factors, such as lipoprotein metabolism or the redistribution of lipids across different organs, rather than organ-specific metabolic activity. This is supported by the significant differences in HDL-TG (HDTG) concentrations between venous comparisons (RRV∼PV, CS∼PV) but not in arteriovenous gradients (A1∼CS, A2∼RRV). These changes may reflect systemic lipid redistribution rather than direct organ-specific uptake or release. Notably, previous studies have demonstrated that arterial lipoprotein (HDL and LDL) and TG concentrations are significantly lower than venous concentrations, which aligns with our results [16].

Our analysis revealed that the significant lipid variations in the A1∼A2 samples were not influenced by chain length or saturation. Interestingly, only the MG species showed distinct concentration differences within the A1∼A2 samples. When comparing the lipids that showed significant changes exclusively in the A1∼A2 samples to those that were significant in both A1∼A2 and CS∼RRV, no pattern in chain length or saturation emerged. This suggests that lipid species altered in both comparisons reflect systemic fluctuations, whereas those differing only in A1∼A2 may reflect reflecting unique metabolic processes in the arterial circulation before reaching the organs. These findings highlight the complexity of lipid dynamics and underscore the necessity of simultaneous arterial and venous sampling for accurately assessing metabolic processes across multiple organs in arteriovenous studies. While simultaneous sampling has been implemented in previous heart or kidney arteriovenous studies [17–20], our results provide direct evidence supporting its necessity. Precision in sampling from organs such as the kidney and heart are critical to avoid misinterpretation and ensure reliable insights into organ-specific metabolic kinetics.

Our study provides novel insights into organ-specific lipid and lipoprotein metabolism, particularly their differential utilisation and release across AVGs in the heart and kidney, which has not been fully explored in previous human arteriovenous studies. Assessing AVGs enabled the capture of organ-specific metabolism within the heart and kidneys, by clarifying whether these organs were taking up or releasing lipids, lipoproteins, or small molecules, providing a direct measure of metabolic exchange at the tissue level. This approach offered insights not reflected in PV sampling, which represents a broader systemic metabolic profile influenced by multiple organs and factors such as diet and hormonal regulation [9]. Our study indicates that the heart preferentially utilises long-chain fatty acids as a metabolic fuel, with slightly negative AVGs (e.g., V1TG, V1CH) suggesting a modest uptake of lipoprotein components. These findings are supported by Voros et al. [19] who reported cardiac uptake of total FA’s, with this uptake significantly increasing in participants with aortic stenosis compared to controls. Similarly, Murashige et al. [18] observed net uptake of FAs independent of chain length or unsaturation, reinforcing the heart’s primary reliance on free fatty acids for energy rather than lipoprotein-derived lipids [21]. In contrast, the kidney sampling reflected an uptake of long-chain FAs with shorter chain lengths (e.g. FA(14:1), FA(16:1), FA(16:2)), which, while relying on the carnitine shuttle for mitochondrial transport, are shorter and less saturated than the very long-chain polyunsaturated fatty acids (e.g. FA(22:4), FA(22:5)) preferentially taken up by the heart [22]. This suggests that the renal lipid metabolism prioritised fatty acids that are more readily oxidised, whilst the heart predominantly relied on substrates that support sustained energy demands. These results align with previous findings in the literature. Kelly et al. [23] reported that kidney AVGs in female mice on a ketogenic diet for five weeks showed uptake of FA(16:0), FA(18:0), FA(18:1), and FA(18:2).

Our analysis reveals a strong negative AVG for acetoacetic acid in the heart, highlighting its role as a key consumer of ketone bodies during fasting or metabolic stress. Supporting this, Avogaro et al. [20] reported significant uptake of ketones, including acetoacetate, in patients with insulin dependent diabetes mellitus when compared to controls. Additionally, the uptake of pyruvate and glutamic acid (which converts to α-ketoglutarate) suggests integration with the TCA cycle, supporting ATP production and further emphasising the heart’s metabolic flexibility and ability to adapt to varying energy substrates [24]. Similar metabolic pathways have been observed in skeletal muscle, where pyruvate and glutamate contribute to α-ketoglutarate production, highlighting a shared mechanism in tissues with high energy demands [25].

In contrast, the kidney demonstrates a positive AVG for acetoacetic acid, indicating active ketogenesis, where fatty acids are oxidised to produce ketone bodies. This aligns with evidence from murine models showing renal expression of mitochondrial hydroxymethylglutaryl-CoA synthase 2 (HMGCS2), the rate-limiting enzyme in ketogenesis, which facilitates fatty acid oxidation to produce ketone bodies to provide energy to peripheral tissues such as the heart [26]. Additionally, our results indicate that the kidney preferentially utilises citrate as an energy source, which is consistent with previous research showing renal arteriovenous concentration differences in porcine models across eight other metabolic organs [15].

The distinct organ-specific handling of lipids, lipoproteins, and small molecules has significant implications for understanding metabolic disturbances in CKM syndrome. Animal models, particularly mice, have been pivotal in elucidating the organ-specific lipid, lipoprotein, and energy metabolism of the heart and kidneys, providing insights that are difficult to obtain from human studies due to ethical and technical constraints [27]. However, key differences exist between species, particularly in lipid and lipoprotein metabolism, as mice primarily transport cholesterol via HDL, whereas humans rely more on low-density lipoproteins (LDL) [20,21]. These differences underscore the importance of human-specific approaches, such as arteriovenous sampling, to directly investigate organ-specific lipid and lipoprotein dynamics.

### Strengths and Limitations

This study is the first to demonstrate that even minor discrepancies in timing between serial arterial sampling (A1 and A2) can alter lipid, lipoprotein and small molecule concentrations. Furthermore, our study uniquely applies arteriovenous sampling from the heart and kidney to reveal organ-specific differences in lipid, lipoprotein, and small molecule handling. While previous research has utilised arteriovenous sampling from the heart and kidneys, focusing angiotensin I and angiotensin II, our study expands on this approach by exploring lipid metabolism [28].

Our study also suffers from some limitations. Samples were collected from participants enrolled in an ongoing randomised, placebo-controlled, crossover trial which remains blinded; hence our results reflect average metabolite gradients across both treatment groups not accounting for the influence of SGLT2 inhibition. Another limitation is that while AVGs measure net metabolite uptake or release across the heart and kidney, they do not account for absolute metabolic flux. Accurate metabolic flux calculations require consideration of blood flow, as vessel diameters vary between the radial artery, CS and RRV [29–32]. Differences in blood flow rates could influence metabolite exchange and clearance, affecting the interpretation of net uptake or release of lipids, lipoproteins and small molecules [29]. Future studies incorporating arteriovenous sampling with blood flow measurements would allow for more precise quantification of metabolic flux, enabling a more precise assessment of organ-specific metabolism.

## Conclusion

This study provides new insights into the interactions of lipids, lipoproteins, and small molecules between the heart and kidneys using arteriovenous sampling and metabolic phenotyping. These findings demonstrate the value of arteriovenous sampling in research on cardiovascular and renal metabolism in individuals with CKM syndrome. The comparison between the heart and kidneys revealed distinct metabolic roles, highlighting the importance of understanding organ-specific characteristics rather than relying on traditional peripheral sampling to determine metabolic alterations.

Arteriovenous sampling combined with comprehensive metabolic phenotyping provides detailed insight into metabolic function at the organ level. This novel approach enhances understanding of metabolic disturbances in CKM syndrome, offering a framework for investigating organ-specific metabolic regulation. While the current study focused on estimating metabolism within individual organs, this approach allows for the future exploration of metabolic crosstalk between organs in control and disease states or in response to therapeutic interventions.

## Data Availability

All data produced in the present study are available upon reasonable request to the authors

