## Supplementary material for "Arteriovenous Sampling for Organ-Specific Metabolic Insights in CKM Syndrome"

**Table S1.** Demographic table of study population at baseline

|  | Female (n=8) | | Male (n=10) | |  |
| --- | --- | --- | --- | --- | --- |
| Variables | **Mean** | **SD** | **Mean** | **SD** | ***p*-value** |
| Age (years) | 51.9 | 14.5 | 57.8 | 8.1 | 0.5 |
| BMI (kg/m^2) | 36.7 | 5.5 | 34.6 | 4.3 | 0.4 |
| Waist (cm) | 110.3 | 10.1 | 116.7 | 10.2 | 0.2 |
| Triglycerides (mmol/L) | 2.2 | 0.7 | 3.1 | 2.7 | 0.9 |
| HDL-C (mmol/L) | 1.1 | 0.2 | 1.0 | 0.2 | 0.4 |
| LDL-C (mmol/L) | 3.3 | 0.7 | 3.2 | 0.8 | 0.6 |
| Glucose (mmol/L) | 7.1 | 3.2 | 5.8 | 0.9 | 0.5 |
| SBP (mmHg) | 124.6 | 11.4 | 137.7 | 15.2 | 0.2 |
| DBP (mmHg) | 81.8 | 6.4 | 84.4 | 10.0 | 0.7 |
| eGFR (mL/min/1.73m^2) | 86.2 | 5.5 | 87.8 | 4.4 | 0.6 |
| *Note*: Demographic statistics include mean and standard deviation. Wilcoxon test p-values indicate the significance of differences between females and males | | | | | |

Box S1. LC-MS/MS analysis method

*Chemical reagents*

Optima^TM^ LC-MS grade acetonitrile (ACN) and isopropanol (IPA) from Thermo Fisher Scientific (Malaga, WA, Australia) with ammonium acetate obtained from Sigma Aldrich (North Ryde, NSW, Australia). A Milli-Q IQ 7000 water purification system (Merck Millipore, Burlington Massachusetts, USA) provided ultrapure LC-MS grade water. Stable isotope labelled internal standards (ISTD) used for LC-MS/MS were Avanti Polar Lipids UltimateSPLASH^TM^ ONE, SphingoSPLASH^TM^ I, 17:1 Lyso PS, 18:1-d7 MG (Sigma-Aldrich, North Ryde, NSW, Australia), Linoleic acid-d11, Arachidonic acid-d5, Palmitic acid-d5, Stearic acid-d4 (Sapphire Bioscience, Redfern, NSW, Australia).

*LC-MS/MS analysis*

Targeted lipid analysis of plasma samples was conducted in a single run using LC-MS/MS with an ExionLC^TM^ system coupled to a QTRAP 6500+ mass spectrometer (SCIEX, Concord, CA), with a 5 µL sample injection volume. To ensure an unbiased analysis, we utilised an in-house R script to randomise sample placement across plates and acquisition order while maintaining all samples from each participant on the same plate. Reversed-phase LC was performed using an Acquity UPLC BEH C18 1.7 µm, 2.1 x100 mm column (Waters Corporation, MA, USA) at 60°C. Mobile phase A was IPA:ACN:H_2_O (20:30:50, v/v/v) containing 10 mM ammonium acetate. Mobile phase B was IPA:ACN:H_2_O (90:9:1, v/v/v) containing 10 mM ammonium acetate. The flow rate was 0.4 mL/min with an initial gradient of 10% B, ramping to 100% B at 12 min, followed by a 2 min re-equilibration period for a total cycle time of 15 min. Key instrument settings included a capillary voltage of +5500 V for positive and −4500 V for negative ion modes, a temperature of 300°C, curtain gas at 20 psi, ion source gas 1 at 40 psi, and ion source gas 2 at 60 psi. Sample plates were stored at 10°C in the autosampler.

Box S2. ^1^H NMR spectroscopic analysis method

*Quantification of Plasma Lipoproteins*

Regression experiments were conducted to quantify 112 parameters of main plasma lipoprotein classes and subclasses using the B.I.LISA method. This involved quantifying the −CH2 (δ = 1.25) and −CH3 (δ = 0.8) peaks of the 1D spectrum after normalisation to the Bruker QuantRef manager within Topspin^TM^ using a partial least squares (PLS-2) regression model. Parameters measured included total plasma lipid analytes such as triglycerides, cholesterol, free cholesterol, phospholipids, apolipoproteins A1/A2/B100, and the B100/A1 ratio, along with their distributions in the different density classes of the plasma lipoproteins. The lipoprotein subclasses included molecular components of VLDL (0.950–1.006 kg/L), LDL (1.09–1.63 kg/L), IDL (1.006–1.019 kg/L), and HDL (1.063–1.210 kg/L). The LDL subfraction was further divided into six density classes (LDL1, 1.019–1.031 kg/L; LDL2, 1.031–1.034 kg/L; LDL3, 1.034–1.037 kg/L; LDL4, 1.037–1.040 kg/L; LDL5, 1.040–1.044 kg/L; and LDL6, 1.044–1.063 kg/L), and the HDL subfractions were categorised into four density classes (HDL1, 1.063–1.100 kg/L; HDL2, 1.100–1.125 kg/L; HDL3, 1.125–1.175 kg/L; and HDL4, 1.175–1.210 kg/L). The full description of the lipoprotein parameters is provided in Table S2.

**Table S2. Annotation of the keys used by the Bruker IVDr Lipoprotein Subclass (B.I.-LISA^TM^) method.** Abbreviations: LDL – low-density lipoprotein; HDL – high-density lipoprotein; VLDL – very low-density lipoprotein; IDL – intermediate-density lipoprotein.

| Key | Lipoprotein | Concentration unit |
| --- | --- | --- |
| TPTG | Triglycerides, total | mg/dL |
| TPCH | Cholesterol, total | mg/dL |
| LDCH | Cholesterol, LDL | mg/dL |
| HDCH | Cholesterol, HDL | mg/dL |
| TPA1 | Apo-A1, total | mg/dL |
| TPA2 | Apo-A2, total | mg/dL |
| TPAB | Apo-B100, total | mg/dL |
| LDHD | LDL-Chol: HDL-Chol | -/- |
| ABA1 | Apo-B100:Apo-A1 | -/- |
| TBPN | Apo-B100, particle number | nmol/L |
| VLPN | VLDL, particle number | nmol/L |
| IDPN | IDL, particle number | nmol/L |
| LDPN | LDL, particle number | nmol/L |
| VLTG | Triglycerides, VLDL | mg/dL |
| IDTG | Triglycerides, IDL | mg/dL |
| LDTG | Triglycerides, LDL | mg/dL |
| HDTG | Triglycerides, HDL | mg/dL |
| VLCH | Cholesterol, VLDL | mg/dL |
| IDCH | Cholesterol, IDL | mg/dL |
| LDCH | Cholesterol, LDL | mg/dL |
| HDCH | Cholesterol, HDL | mg/dL |
| VLFC | Free Cholesterol, VLDL | mg/dL |
| IDFC | Free Cholesterol, IDL | mg/dL |
| LDFC | Free Cholesterol, LDL | mg/dL |
| HDFC | Free Cholesterol, HDL | mg/dL |
| VLPL | Phospholipids, VLDL | mg/dL |
| IDPL | Phospholipids, IDL | mg/dL |
| LDPL | Phospholipids, LDL | mg/dL |
| HDPL | Phospholipids, HDL | mg/dL |
| HDA1 | Apo-A1, HDL | mg/dL |
| HDA2 | Apo-A2, HDL | mg/dL |
| VLAB | Apo-B, VLDL | mg/dL |
| IDAB | Apo-B, IDL | mg/dL |
| LDAB | Apo-B, LDL | mg/dL |
| VLDL Subfractions | | |
| V1TG | Triglycerides, VLDL-1 | mg/dL |
| V2TG | Triglycerides, VLDL-2 | mg/dL |
| V3TG | Triglycerides, VLDL-3 | mg/dL |
| V4TG | Triglycerides, VLDL-4 | mg/dL |
| V5TG | Triglycerides, VLDL-5 | mg/dL |
| V1CH | Cholesterol, VLDL-1 | mg/dL |
| V2CH | Cholesterol, VLDL-2 | mg/dL |
| V3CH | Cholesterol, VLDL-3 | mg/dL |
| V4CH | Cholesterol, VLDL-4 | mg/dL |
| V5CH | Cholesterol, VLDL-5 | mg/dL |
| V1FC | FreeCholesterol, VLDL-1 | mg/dL |
| V2FC | FreeCholesterol, VLDL-2 | mg/dL |
| V3FC | FreeCholesterol, VLDL-3 | mg/dL |
| V4FC | FreeCholesterol, VLDL-4 | mg/dL |
| V5FC | FreeCholesterol, VLDL-5 | mg/dL |
| V1PL | Phospholipids, VLDL-1 | mg/dL |
| V2PL | Phospholipids, VLDL-2 | mg/dL |
| V3PL | Phospholipids, VLDL-3 | mg/dL |
| V4PL | Phospholipids, VLDL-4 | mg/dL |
| V5PL | Phospholipids, VLDL-5 | mg/dL |
| LDL Subfractions | | |
| L1PN | LDL1, particle number | nmol/L |
| L2PN | LDL2, particle number | nmol/L |
| L3PN | LDL3, particle number | nmol/L |
| L4PN | LDL4, particle number | nmol/L |
| L5PN | LDL5, particle number | nmol/L |
| L6PN | LDL6, particle number | nmol/L |
| L1TG | Triglycerides, LDL-1 | mg/dL |
| L2TG | Triglycerides, LDL-2 | mg/dL |
| L3TG | Triglycerides, LDL-3 | mg/dL |
| L4TG | Triglycerides, LDL-4 | mg/dL |
| L5TG | Triglycerides, LDL-5 | mg/dL |
| L6TG | Triglycerides, LDL-6 | mg/dL |
| L1CH | Cholesterol, LDL-1 | mg/dL |
| L2CH | Cholesterol, LDL-2 | mg/dL |
| L3CH | Cholesterol, LDL-3 | mg/dL |
| L4CH | Cholesterol, LDL-4 | mg/dL |
| L5CH | Cholesterol, LDL-5 | mg/dL |
| L6CH | Cholesterol, LDL-6 | mg/dL |
| L1FC | FreeCholesterol, LDL-1 | mg/dL |
| L2FC | FreeCholesterol, LDL-2 | mg/dL |
| L3FC | FreeCholesterol, LDL-3 | mg/dL |
| L4FC | FreeCholesterol, LDL-4 | mg/dL |
| L5FC | FreeCholesterol, LDL-5 | mg/dL |
| L6FC | FreeCholesterol, LDL-6 | mg/Dl |
| L1PL | Phospholipids, LDL-1 | mg/dL |
| L2PL | Phospholipids, LDL-2 | mg/dL |
| L3PL | Phospholipids, LDL-3 | mg/dL |
| L4PL | Phospholipids, LDL-4 | mg/dL |
| L5PL | Phospholipids, LDL-5 | mg/dL |
| L1PL | Phospholipids, LDL-6 | mg/dL |
| L1AB | Apo-B, LDL-1 | mg/dL |
| L2AB | Apo-B, LDL-2 | mg/dL |
| L3AB | Apo-B, LDL-3 | mg/dL |
| L4AB | Apo-B, LDL-4 | mg/dL |
| L5AB | Apo-B, LDL-5 | mg/dL |
| L6AB | Apo-B, LDL-6 | mg/dL |
| HDL Subfractions | | |
| H1TG | Triglycerides, HDL-1 | mg/dL |
| H2TG | Triglycerides, HDL-2 | mg/dL |
| H3TG | Triglycerides, HDL-3 | mg/dL |
| H4TG | Triglycerides, HDL-4 | mg/dL |
| H1CH | Cholesterol, HDL-1 | mg/dL |
| H2CH | Cholesterol, HDL-2 | mg/dL |
| H3CH | Cholesterol, HDL-3 | mg/dL |
| H4CH | Cholesterol, HDL-4 | mg/dL |
| H1FC | Free Cholesterol, HDL-1 | mg/dL |
| H2FC | Free Cholesterol, HDL-2 | mg/dL |
| H3FC | Free Cholesterol, HDL-3 | mg/dL |
| H4FC | Free Cholesterol, HDL-4 | mg/dL |
| H1PL | Phospholipids, HDL-1 | mg/dL |
| H2PL | Phospholipids, HDL-2 | mg/dL |
| H3PL | Phospholipids, HDL-3 | mg/dL |
| H4PL | Phospholipids, HDL-4 | mg/dL |
| H1A1 | Apo-A1, HDL-1 | mg/dL |
| H2A1 | Apo-A1, HDL-2 | mg/dL |
| H3A1 | Apo-A1, HDL-3 | mg/dL |
| H4A1 | Apo-A1, HDL-4 | mg/dL |
| H1A2 | Apo-A2, HDL-1 | mg/dL |
| H2A2 | Apo-A2, HDL-2 | mg/dL |
| H3A2 | Apo-A2, HDL-3 | mg/dL |
| H4A2 | Apo-A2, HDL-4 | mg/dL |

**Table S3. Annotation of the low molecular weight metabolites used by the Bruker IVDr B.I. Quant-PS^TM^ method**

| Metabolite | Concentration unit |
| --- | --- |
| Acetic acid | mmol/L |
| Acetoacetic acid | mmol/L |
| Acetone | mmol/L |
| Alanine | mmol/L |
| Citric acid | mmol/L |
| Creatine | mmol/L |
| Formic acid | mmol/L |
| Glucose | mmol/L |
| Glutamic acid | mmol/L |
| Glutamine | mmol/L |
| Glycine | mmol/L |
| Histidine | mmol/L |
| D-3-hydroxybutyric acid | mmol/L |
| Isoleucine | mmol/L |
| Lactic acid | mmol/L |
| Leucine | mmol/L |
| Lysine | mmol/L |
| N,N-dimethylglycine | mmol/L |
| Methionine | mmol/L |
| Phenylalanine | mmol/L |
| Pyruvic acid | mmol/L |
| Trimethylamine-N-oxide | mmol/L |
| Tyrosine | mmol/L |
| Valine | mmol/L |

**Table S4 Significant lipids with an adjusted *p*-value < 0.05 from pairwise Wilcoxon signed-rank test.** Significance denoted by * = *p* < 0.05, ** = *p* < 0.01, *** = *p* < 0.001. **↓	↑**

|  | | Sampling site comparison adjusted *p*-value | | | | | |
| --- | --- | --- | --- | --- | --- | --- | --- |
| Lipid | **Subclass** | **A1 ~ A2** | **A1 ~ CS** | **A2 ~ RRV** | **CS ~ RRV** | **RRV ~ PV** | **CS ~ PV** |
| CE(14:0) | CE | n.s. | n.s. | n.s. | n.s. | *** **↑** | *** **↑** |
| CE(18:2) | CE | n.s. | n.s. | n.s. | n.s. | *** **↑** | *** **↑** |
| CE(22:5) | CE | n.s. | n.s. | n.s. | n.s. | *** **↓** | *** **↓** |
| CE(22:6) | CE | n.s. | n.s. | n.s. | n.s. | *** **↓** | *** **↓** |
| DG(14:0_18:1) | DG | n.s. | n.s. | n.s. | n.s. | * **↑** | n.s. |
| DG(14:0_20:4) | DG | n.s. | n.s. | * **↓** | n.s. | ** **↓** | *** **↓** |
| DG(16:0_22:6) | DG | n.s. | n.s. | n.s. | n.s. | * **↓** | ** **↓** |
| DG(16:1_20:4) | DG | n.s. | n.s. | n.s. | n.s. | *** **↓** | *** **↓** |
| DG(16:1_22:6) | DG | n.s. | n.s. | n.s. | n.s. | n.s. | ** **↓** |
| DG(18:0_18:3) | DG | n.s. | n.s. | n.s. | n.s. | * **↑** | * **↑** |
| DG(18:1_20:3) | DG | n.s. | n.s. | n.s. | * **↑** | *** **↓** | *** **↓** |
| DG(18:1_20:4) | DG | n.s. | n.s. | n.s. | n.s. | *** **↓** | *** **↓** |
| DG(18:1_20:5) | DG | n.s. | n.s. | n.s. | n.s. | *** **↓** | *** **↓** |
| DG(18:1_22:5) | DG | n.s. | n.s. | n.s. | n.s. | * **↓** | * **↓** |
| DG(18:1_22:6) | DG | n.s. | n.s. | n.s. | n.s. | *** **↓** | *** **↓** |
| DG(18:2_20:3) | DG | n.s. | n.s. | n.s. | n.s. | *** **↓** | *** **↓** |
| DG(18:2_20:4) | DG | * **↑** | n.s. | n.s. | * **↓** | *** **↓** | ** **↓** |
| DG(18:2_20:5) | DG | n.s. | n.s. | n.s. | n.s. | *** **↓** | *** **↓** |
| DG(18:2_22:5) | DG | n.s. | n.s. | n.s. | n.s. | n.s. | ** **↓** |
| DG(18:2_22:6) | DG | n.s. | n.s. | n.s. | n.s. | ** **↓** | ** **↓** |
| DG(20:0_20:0) | DG | n.s. | n.s. | n.s. | n.s. | *** **↑** | *** **↑** |
| FA(14:0) | FA | n.s. | n.s. | *** **↓** | * **↑** | *** **↓** | ** **↓** |
| FA(14:1) | FA | * **↑** | n.s. | ** **↓** | n.s. | *** **↓** | *** **↓** |
| FA(16:0) | FA | ** **↑** | n.s. | n.s. | * **↑** | *** **↓** | *** **↓** |
| FA(16:1) | FA | ** **↑** | n.s. | * **↓** | * **↑** | *** **↓** | *** **↓** |
| FA(16:2) | FA | ** **↑** | n.s. | * **↓** | n.s. | *** **↓** | *** **↓** |
| FA(18:0) | FA | n.s. | * **↓** | n.s. | ** **↑** | *** **↓** | *** **↓** |
| FA(18:1) | FA | n.s. | * **↓** | n.s. | * **↑** | *** **↓** | *** **↓** |
| FA(18:2) | FA | ** **↑** | n.s. | n.s. | ** **↑** | *** **↓** | *** **↓** |
| FA(18:3) | FA | * **↑** | n.s. | * **↓** | * **↑** | *** **↓** | *** **↓** |
| FA(20:0) | FA | ** **↑** | * **↓** | n.s. | * **↑** | *** **↓** | *** **↓** |
| FA(20:1) | FA | ** **↑** | *** **↓** | n.s. | *** **↑** | *** **↓** | *** **↓** |
| FA(20:2) | FA | ** **↑** | *** **↓** | n.s. | *** **↑** | *** **↓** | *** **↓** |
| FA(20:3) | FA | ** **↑** | * **↓** | n.s. | ** **↑** | *** **↓** | *** **↓** |
| FA(20:4) | FA | *** **↑** | n.s. | n.s. | ** **↑** | *** **↓** | *** **↓** |
| FA(20:5) | FA | ** **↑** | n.s. | n.s. | ** **↑** | *** **↓** | *** **↓** |
| FA(22:4) | FA | ** **↑** | *** **↓** | n.s. | ** **↑** | *** **↓** | *** **↓** |
| FA(22:5) | FA | ** **↑** | * **↓** | n.s. | ** **↑** | *** **↓** | *** **↓** |
| FA(22:6) | FA | ** **↑** | n.s. | n.s. | * **↑** | *** **↓** | *** **↓** |
| FA(24:0) | FA | n.s. | n.s. | n.s. | n.s. | *** **↓** | n.s. |
| FA(24:1) | FA | ** **↑** | n.s. | n.s. | * **↑** | *** **↓** | n.s. |
| LPC(18:1) | LPC | ** **↑** | n.s. | n.s. | ** **↑** | *** **↓** | ** **↓** |
| LPC(18:2) | LPC | ** **↑** | n.s. | n.s. | ** **↑** | *** **↓** | *** **↓** |
| LPC(18:3) | LPC | ** **↑** | n.s. | n.s. | n.s. | *** **↓** | *** **↓** |
| LPC(20:2) | LPC | ** **↑** | n.s. | n.s. | n.s. | *** **↓** | *** **↓** |
| LPC(20:3) | LPC | *** **↑** | n.s. | n.s. | n.s. | *** **↓** | *** **↓** |
| LPC(20:4) | LPC | *** **↑** | n.s. | n.s. | n.s. | *** **↓** | *** **↓** |
| LPC(20:5) | LPC | *** **↑** | n.s. | n.s. | * **↑** | *** **↓** | *** **↓** |
| LPC(22:4) | LPC | *** **↑** | n.s. | n.s. | * **↑** | *** **↓** | *** **↓** |
| LPC(22:5) | LPC | ** **↑** | n.s. | n.s. | n.s. | *** **↓** | *** **↓** |
| LPC(22:6) | LPC | ** **↑** | n.s. | n.s. | n.s. | *** **↓** | *** **↓** |
| LPE(16:1) | LPE | n.s. | n.s. | n.s. | n.s. | *** **↓** | *** **↓** |
| LPE(18:0) | LPE | n.s. | n.s. | n.s. | n.s. | *** **↑** | *** **↑** |
| LPE(18:1) | LPE | * **↑** | n.s. | n.s. | n.s. | *** **↓** | *** **↓** |
| LPE(18:2) | LPE | ** **↑** | * **↑** | n.s. | n.s. | *** **↓** | *** **↓** |
| LPE(18:3) | LPE | ** **↑** | n.s. | n.s. | n.s. | *** **↓** | ** **↓** |
| LPE(20:0) | LPE | *** **↑** | n.s. | n.s. | ** **↑** | *** **↓** | ** **↓** |
| LPE(20:1) | LPE | * **↑** | n.s. | n.s. | * **↑** | *** **↓** | * **↓** |
| LPE(20:2) | LPE | ** **↑** | n.s. | n.s. | * **↑** | *** **↓** | *** **↓** |
| LPE(20:3) | LPE | ** **↑** | n.s. | n.s. | n.s. | *** **↓** | *** **↓** |
| LPE(20:4) | LPE | ** **↑** | n.s. | * **↑** | n.s. | *** **↓** | *** **↓** |
| LPE(20:5) | LPE | n.s. | n.s. | n.s. | n.s. | *** **↓** | *** **↓** |
| LPE(22:4) | LPE | ** **↑** | n.s. | n.s. | ** **↑** | *** **↓** | *** **↓** |
| LPE(22:5) | LPE | ** **↑** | n.s. | n.s. | * **↑** | *** **↓** | *** **↓** |
| LPE(22:6) | LPE | * **↑** | n.s. | * **↓** | n.s. | *** **↓** | *** **↓** |
| LPG(18:0) | LPG | * **↑** | n.s. | n.s. | * **↑** | * **↓** | n.s. |
| LPG(18:1) | LPG | * **↑** | n.s. | n.s. | n.s. | *** **↓** | *** **↓** |
| LPG(18:2) | LPG | ** **↑** | n.s. | n.s. | n.s. | *** **↓** | *** **↓** |
| LPG(20:1) | LPG | n.s. | n.s. | n.s. | ** **↑** | *** **↓** | ** **↓** |
| LPG(22:4) | LPG | ** **↑** | n.s. | n.s. | ** **↑** | *** **↓** | ** **↓** |
| LPG(22:5) | LPG | *** **↑** | n.s. | n.s. | ** **↑** | *** **↓** | *** **↓** |
| LPI(16:1) | LPI | ** **↑** | n.s. | * **↓** | n.s. | *** **↓** | *** **↓** |
| LPI(18:1) | LPI | n.s. | n.s. | n.s. | n.s. | *** **↓** | *** **↓** |
| LPI(18:2) | LPI | *** **↑** | n.s. | n.s. | n.s. | *** **↓** | *** **↓** |
| LPI(20:0) | LPI | * **↑** | n.s. | n.s. | *** **↑** | ** **↓** | *** **↓** |
| LPI(20:2) | LPI | n.s. | n.s. | n.s. | n.s. | *****↓** | ** **↓** |
| LPI(20:3) | LPI | *** **↑** | n.s. | n.s. | * **↑** | *** **↓** | *** **↓** |
| LPI(20:4) | LPI | *** **↑** | n.s. | n.s. | * **↑** | *** **↓** | *** **↓** |
| LPS(16:0) | LPS | ** **↑** | n.s. | n.s. | n.s. | *** **↓** | ** **↓** |
| LPS(18:0) | LPS | ** **↑** | n.s. | n.s. | *** **↑** | *** **↓** | *** **↓** |
| LPS(18:1) | LPS | ** **↑** | n.s. | n.s. | *** **↑** | *** **↓** | ** **↓** |
| LPS(20:1) | LPS | n.s. | n.s. | n.s. | n.s. | *** **↓** | * **↓** |
| LPS(20:2) | LPS | n.s. | n.s. | n.s. | * **↑** | *** **↓** | n.s. |
| LPS(20:3) | LPS | n.s. | n.s. | n.s. | * **↑** | *** **↓** | n.s. |
| LPS(20:4) | LPS | *** **↑** | n.s. | n.s. | * **↑** | *** **↓** | ** **↓** |
| MG(14:0) | MG | n.s. | n.s. | n.s. | n.s. | *** **↓** | *** **↓** |
| MG(16:0) | MG | ** **↑** | n.s. | n.s. | n.s. | *** **↓** | *** **↓** |
| MG(16:1) | MG | ** **↑** | n.s. | n.s. | n.s. | *** **↓** | *** **↓** |
| MG(18:0) | MG | * **↑** | n.s. | n.s. | n.s. | *** **↓** | *** **↓** |
| MG(18:1) | MG | ** **↑** | n.s. | n.s. | n.s. | *** **↓** | *** **↓** |
| MG(18:2) | MG | *** **↑** | n.s. | n.s. | n.s. | *** **↓** | *** **↓** |
| MG(18:3) | MG | ** **↑** | n.s. | n.s. | n.s. | *** **↓** | *** **↓** |
| MG(20:3) | MG | *** **↑** | n.s. | n.s. | n.s. | *** **↓** | *** **↓** |
| MG(20:4) | MG | ** **↑** | n.s. | n.s. | n.s. | *** **↓** | *** **↓** |
| MG(22:3) | MG | * **↑** | n.s. | n.s. | * **↑** | *** **↓** | *** **↓** |
| MG(22:4) | MG | ** **↑** | n.s. | n.s. | n.s. | *** **↓** | *** **↓** |
| MG(22:5) | MG | *** **↑** | n.s. | n.s. | n.s. | *** **↓** | *** **↓** |
| MG(22:6) | MG | *** **↑** | n.s. | n.s. | n.s. | *** **↓** | *** **↓** |
| PE(18:0_18:0) | PE | n.s. | n.s. | n.s. | n.s. | *** **↓** | *** **↓** |
| PE(18:0_18:1) | PE | n.s. | n.s. | n.s. | n.s. | *** **↓** | *** **↓** |
| PE(18:0_18:2) | PE | * **↓** | n.s. | n.s. | n.s. | *** **↑** | *** **↑** |
| PE(18:0_22:6) | PE | n.s. | n.s. | n.s. | n.s. | *** **↑** | *** **↑** |
| PE(18:1_18:1) | PE | n.s. | n.s. | n.s. | n.s. | *** **↑** | *** **↑** |
| PE(18:1_22:4) | PE | n.s. | n.s. | n.s. | n.s. | *** **↑** | *** **↑** |
| PE(18:1_22:5) | PE | n.s. | n.s. | n.s. | n.s. | *** **↑** | *** **↑** |
| PE(O-18:0_18:2) | PE | n.s. | n.s. | n.s. | n.s. | *** **↑** | *** **↑** |
| PE(O-18:0_22:4) | PE | n.s. | n.s. | n.s. | n.s. | *** **↑** | *** **↑** |
| PG(16:0_18:1) | PG | n.s. | n.s. | n.s. | n.s. | *** **↑** | *** **↑** |
| PG(18:0_18:1) | PG | n.s. | n.s. | n.s. | n.s. | *** **↑** | *** **↑** |
| PG(18:0_18:2) | PG | n.s. | n.s. | n.s. | n.s. | *** **↑** | *** **↑** |
| PS(14:1_14:1) | PS | * **↑** | n.s. | n.s. | *** **↑** | *** **↓** | * **↓** |
| PS(18:0_18:1) | PS | n.s. | n.s. | n.s. | n.s. | *** **↑** | *** **↑** |
| PS(18:0_20:1) | PS | n.s. | n.s. | n.s. | n.s. | *** **↑** | ** **↑** |
| PS(18:1_18:2) | PS | n.s. | n.s. | n.s. | n.s. | *** **↑** | *** **↑** |
| TG(44:3_FA18:2) | TG | * **↓** | n.s. | n.s. | ** **↓** | *** **↑** | *** **↑** |
| TG(46:1_FA18:1) | TG | ** **↓** | n.s. | n.s. | ** **↓** | *** **↑** | *** **↑** |
| TG(46:2_FA14:0) | TG | * **↓** | n.s. | n.s. | * **↓** | *** **↑** | * **↑** |
| TG(46:2_FA18:1) | TG | * **↓** | n.s. | n.s. | * **↓** | *** **↑** | *** **↑** |
| TG(46:2_FA18:2) | TG | * **↓** | n.s. | n.s. | ** **↓** | *** **↑** | *** **↑** |
| TG(46:3_FA14:0) | TG | * **↓** | n.s. | n.s. | * **↓** | *** **↑** | *** **↑** |
| TG(46:3_FA18:2) | TG | * **↓** | n.s. | n.s. | ** **↓** | *** **↑** | *** **↑** |
| TG(47:1_FA14:0) | TG | * **↓** | n.s. | n.s. | ** **↓** | *** **↑** | *** **↑** |
| TG(47:2_FA14:0) | TG | n.s. | n.s. | n.s. | * **↓** | *** **↑** | *** **↑** |
| TG(47:2_FA18:1) | TG | * **↓** | n.s. | n.s. | * **↓** | *** **↑** | *** **↑** |
| TG(47:2_FA18:2) | TG | n.s. | n.s. | n.s. | * **↓** | *** **↑** | *** **↑** |
| TG(48:1_FA14:0) | TG | ** **↓** | n.s. | n.s. | ** **↓** | *** **↑** | *** **↑** |
| TG(48:1_FA16:0) | TG | ** **↓** | n.s. | n.s. | ** **↓** | *** **↑** | *** **↑** |
| TG(48:1_FA18:1) | TG | ** **↓** | n.s. | n.s. | ** **↓** | *** **↑** | *** **↑** |
| TG(48:2_FA14:0) | TG | * **↓** | n.s. | n.s. | * **↓** | *** **↑** | *** **↑** |
| TG(48:2_FA16:0) | TG | * **↓** | n.s. | n.s. | * **↓** | *** **↑** | *** **↑** |
| TG(48:2_FA18:1) | TG | * **↓** | n.s. | n.s. | * **↓** | *** **↑** | *** **↑** |
| TG(48:2_FA18:2) | TG | * **↓** | n.s. | n.s. | * **↓** | *** **↑** | *** **↑** |
| TG(48:3_FA14:0) | TG | * **↓** | n.s. | n.s. | n.s. | *** **↑** | *** **↑** |
| TG(48:3_FA16:0) | TG | * **↓** | n.s. | n.s. | n.s. | *** **↑** | *** **↑** |
| TG(48:3_FA16:1) | TG | * **↓** | n.s. | n.s. | * **↓** | *** **↑** | *** **↑** |
| TG(48:3_FA18:1) | TG | * **↓** | n.s. | n.s. | ** **↓** | *** **↑** | *** **↑** |
| TG(48:3_FA18:2) | TG | * **↓** | n.s. | n.s. | * **↓** | *** **↑** | *** **↑** |
| TG(48:3_FA18:3) | TG | n.s. | n.s. | n.s. | * **↓** | *** **↑** | *** **↑** |
| TG(48:4_FA14:0) | TG | n.s. | n.s. | n.s. | * **↓** | *** **↑** | *** **↑** |
| TG(48:4_FA16:0) | TG | n.s. | n.s. | n.s. | * **↓** | *** **↑** | * **↑** |
| TG(48:4_FA16:1) | TG | n.s. | n.s. | n.s. | * **↓** | *** **↑** | *** **↑** |
| TG(48:4_FA18:1) | TG | n.s. | n.s. | n.s. | ** **↓** | *** **↑** | *** **↑** |
| TG(48:4_FA18:2) | TG | * **↓** | n.s. | n.s. | ** **↓** | *** **↑** | *** **↑** |
| TG(48:4_FA18:3) | TG | n.s. | n.s. | n.s. | * **↓** | *** **↑** | *** **↑** |
| TG(48:4_FA20:4) | TG | n.s. | n.s. | n.s. | * **↓** | *** **↑** | *** **↑** |
| TG(48:5_FA18:2) | TG | * **↓** | n.s. | n.s. | ** **↓** | *** **↑** | *** **↑** |
| TG(48:5_FA18:3) | TG | * **↓** | n.s. | n.s. | ** **↓** | *** **↑** | *** **↑** |
| TG(49:0_FA16:0) | TG | * **↓** | n.s. | n.s. | * **↓** | *** **↑** | *** **↑** |
| TG(49:0_FA17:0) | TG | * **↓** | n.s. | n.s. | * **↓** | *** **↑** | *** **↑** |
| TG(49:0_FA18:0) | TG | * **↓** | n.s. | n.s. | * **↓** | *** **↑** | *** **↑** |
| TG(49:1_FA14:0) | TG | * **↓** | n.s. | n.s. | ** **↓** | *** **↑** | *** **↑** |
| TG(49:1_FA16:0) | TG | * **↓** | n.s. | n.s. | * **↓** | *** **↑** | *** **↑** |
| TG(49:1_FA16:1) | TG | * **↓** | n.s. | n.s. | * **↓** | *** **↑** | *** **↑** |
| TG(49:1_FA17:0) | TG | * **↓** | n.s. | n.s. | * **↓** | *** **↑** | *** **↑** |
| TG(49:1_FA18:1) | TG | * **↓** | n.s. | n.s. | ** **↓** | *** **↑** | *** **↑** |
| TG(49:2_FA14:0) | TG | * **↓** | n.s. | n.s. | ** **↓** | *** **↑** | *** **↑** |
| TG(49:2_FA16:0) | TG | * **↓** | n.s. | n.s. | ** **↓** | *** **↑** | *** **↑** |
| TG(49:2_FA16:1) | TG | * **↓** | n.s. | n.s. | ** **↓** | *** **↑** | *** **↑** |
| TG(49:2_FA17:0) | TG | * **↓** | n.s. | n.s. | ** **↓** | *** **↑** | *** **↑** |
| TG(49:2_FA18:1) | TG | * **↓** | n.s. | n.s. | ** **↓** | *** **↑** | *** **↑** |
| TG(49:2_FA18:2) | TG | * **↓** | n.s. | n.s. | ** **↓** | *** **↑** | *** **↑** |
| TG(49:3_FA16:0) | TG | * **↓** | n.s. | n.s. | ** **↓** | *** **↑** | *** **↑** |
| TG(49:3_FA16:1) | TG | * **↓** | n.s. | n.s. | ** **↓** | *** **↑** | *** **↑** |
| TG(49:3_FA18:2) | TG | * **↓** | n.s. | n.s. | ** **↓** | *** **↑** | *** **↑** |
| TG(49:3_FA18:3) | TG | * **↓** | n.s. | n.s. | * **↓** | *** **↑** | *** **↑** |
| TG(50:0_FA14:0) | TG | * **↓** | n.s. | n.s. | * **↓** | *** **↑** | *** **↑** |
| TG(50:0_FA18:0) | TG | n.s. | n.s. | n.s. | * **↓** | *** **↑** | n.s. |
| TG(50:1_FA14:0) | TG | * **↓** | n.s. | n.s. | ** **↓** | *****↑** | *** **↑** |
| TG(50:1_FA16:0) | TG | * **↓** | n.s. | n.s. | * **↓** | *** **↑** | *** **↑** |
| TG(50:1_FA16:1) | TG | ** **↓** | n.s. | n.s. | * **↓** | *** **↑** | *** **↑** |
| TG(50:1_FA18:0) | TG | ** **↓** | n.s. | n.s. | ** **↓** | *** **↑** | *** **↑** |
| TG(50:1_FA18:1) | TG | ** **↓** | n.s. | n.s. | ** **↓** | *** **↑** | *** **↑** |
| TG(50:1_FA20:1) | TG | *** **↓** | n.s. | n.s. | *** **↓** | *** **↑** | *** **↑** |
| TG(50:2_FA14:0) | TG | * **↓** | n.s. | n.s. | *** **↓** | *** **↑** | *** **↑** |
| TG(50:2_FA16:0) | TG | * **↓** | n.s. | n.s. | * **↓** | *** **↑** | *** **↑** |
| TG(50:2_FA16:1) | TG | * **↓** | n.s. | n.s. | * **↓** | *** **↑** | *** **↑** |
| TG(50:2_FA18:0) | TG | * **↓** | n.s. | n.s. | ** **↓** | *** **↑** | *** **↑** |
| TG(50:2_FA18:1) | TG | n.s. | n.s. | n.s. | * **↓** | *** **↑** | *** **↑** |
| TG(50:2_FA18:2) | TG | * **↓** | n.s. | n.s. | ** **↓** | *** **↑** | *** **↑** |
| TG(50:2_FA20:2) | TG | * **↓** | n.s. | n.s. | * **↓** | *** **↑** | *** **↑** |
| TG(50:3_FA14:0) | TG | * **↓** | n.s. | n.s. | ** **↓** | *** **↑** | *** **↑** |
| TG(50:3_FA16:0) | TG | * **↓** | n.s. | n.s. | ** **↓** | *** **↑** | *** **↑** |
| TG(50:3_FA16:1) | TG | ** **↓** | n.s. | n.s. | ** **↓** | *** **↑** | *** **↑** |
| TG(50:3_FA18:0) | TG | * **↓** | n.s. | n.s. | ** **↓** | *** **↑** | *** **↑** |
| TG(50:3_FA18:1) | TG | ** **↓** | n.s. | n.s. | ** **↓** | *** **↑** | *** **↑** |
| TG(50:3_FA18:2) | TG | * **↓** | n.s. | n.s. | ** **↓** | *** **↑** | *** **↑** |
| TG(50:3_FA18:3) | TG | ** **↓** | n.s. | n.s. | * **↓** | *** **↑** | *** **↑** |
| TG(50:3_FA20:3) | TG | * **↓** | n.s. | n.s. | * **↓** | *** **↑** | *** **↑** |
| TG(50:4_FA14:0) | TG | * **↓** | n.s. | n.s. | ** **↓** | *** **↑** | *** **↑** |
| TG(50:4_FA16:0) | TG | * **↓** | n.s. | n.s. | * **↓** | *** **↑** | *** **↑** |
| TG(50:4_FA16:1) | TG | * **↓** | n.s. | n.s. | * **↓** | *** **↑** | *** **↑** |
| TG(50:4_FA18:1) | TG | * **↓** | n.s. | n.s. | * **↓** | *** **↑** | *** **↑** |
| TG(50:4_FA18:2) | TG | * **↓** | n.s. | n.s. | * **↓** | *** **↑** | *** **↑** |
| TG(50:4_FA18:3) | TG | * **↓** | n.s. | n.s. | * **↓** | *** **↑** | *** **↑** |
| TG(50:4_FA20:3) | TG | n.s. | n.s. | n.s. | * **↓** | *** **↑** | *** **↑** |
| TG(50:4_FA20:4) | TG | n.s. | n.s. | n.s. | ***↓** | *** **↑** | *** **↑** |
| TG(50:5_FA14:0) | TG | * **↓** | n.s. | n.s. | * **↓** | *** **↑** | *** **↑** |
| TG(50:5_FA16:0) | TG | * **↓** | n.s. | n.s. | * **↓** | *** **↑** | *** **↑** |
| TG(50:5_FA16:1) | TG | * **↓** | n.s. | n.s. | * **↓** | *** **↑** | *** **↑** |
| TG(50:5_FA18:1) | TG | * **↓** | n.s. | n.s. | * **↓** | *** **↑** | *** **↑** |
| TG(50:5_FA18:2) | TG | n.s. | n.s. | n.s. | ** **↓** | *****↑** | *** **↑** |
| TG(50:5_FA18:3) | TG | n.s. | n.s. | n.s. | * **↓** | *** **↑** | *** **↑** |
| TG(50:5_FA20:4) | TG | n.s. | n.s. | n.s. | n.s. | *** **↑** | *** **↑** |
| TG(50:5_FA20:5) | TG | n.s. | n.s. | n.s. | n.s. | *** **↑** | *** **↑** |
| TG(50:6_FA20:4) | TG | n.s. | n.s. | n.s. | * **↓** | *** **↑** | *** **↑** |
| TG(51:0_FA16:0) | TG | * **↓** | n.s. | n.s. | ** **↓** | *** **↑** | *** **↑** |
| TG(51:0_FA17:0) | TG | * **↓** | n.s. | n.s. | ** **↓** | *** **↑** | *** **↑** |
| TG(51:0_FA18:0) | TG | * **↓** | n.s. | n.s. | ** **↓** | *** **↑** | *** **↑** |
| TG(51:1_FA16:0) | TG | * **↓** | n.s. | n.s. | ** **↓** | *** **↑** | *** **↑** |
| TG(51:1_FA17:0) | TG | * **↓** | n.s. | n.s. | * **↓** | *** **↑** | *** **↑** |
| TG(51:1_FA18:0) | TG | * **↓** | n.s. | n.s. | * **↓** | *** **↑** | *** **↑** |
| TG(51:1_FA18:1) | TG | * **↓** | n.s. | n.s. | ** **↓** | *** **↑** | *** **↑** |
| TG(51:2_FA16:0) | TG | * **↓** | n.s. | n.s. | * **↓** | *** **↑** | *** **↑** |
| TG(51:2_FA16:1) | TG | * **↓** | n.s. | n.s. | * **↓** | *** **↑** | *** **↑** |
| TG(51:2_FA17:0) | TG | * **↓** | n.s. | n.s. | * **↓** | *** **↑** | *** **↑** |
| TG(51:2_FA18:1) | TG | n.s. | n.s. | n.s. | ** **↓** | *** **↑** | *** **↑** |
| TG(51:2_FA18:2) | TG | * **↓** | n.s. | n.s. | * **↓** | *** **↑** | *** **↑** |
| TG(51:3_FA16:1) | TG | * **↓** | n.s. | n.s. | ** **↓** | *** **↑** | *** **↑** |
| TG(51:3_FA17:0) | TG | * **↓** | n.s. | n.s. | ** **↓** | *** **↑** | *** **↑** |
| TG(51:3_FA18:2) | TG | * **↓** | n.s. | n.s. | ** **↓** | *** **↑** | *** **↑** |
| TG(51:3_FA18:3) | TG | * **↓** | n.s. | n.s. | ** **↓** | *** **↑** | *** **↑** |
| TG(51:4_FA16:1) | TG | * **↓** | n.s. | n.s. | * **↓** | *** **↑** | *** **↑** |
| TG(51:4_FA18:2) | TG | * **↓** | n.s. | n.s. | ** **↓** | *** **↑** | *** **↑** |
| TG(51:4_FA18:3) | TG | * **↓** | n.s. | n.s. | ** **↓** | *** **↑** | *** **↑** |
| TG(51:4_FA20:4) | TG | n.s. | n.s. | n.s. | * **↓** | *** **↑** | *** **↑** |
| TG(51:5_FA18:2) | TG | * **↓** | n.s. | n.s. | ** **↓** | *** **↑** | *** **↑** |
| TG(51:5_FA18:3) | TG | * **↓** | n.s. | n.s. | ** **↓** | *** **↑** | *** **↑** |
| TG(52:0_FA16:0) | TG | * **↓** | n.s. | n.s. | ** **↓** | *** **↑** | *** **↑** |
| TG(52:0_FA18:0) | TG | ** **↓** | n.s. | n.s. | ** **↓** | *** **↑** | *** **↑** |
| TG(52:0_FA20:0) | TG | n.s. | n.s. | n.s. | * **↓** | *** **↑** | *** **↑** |
| TG(52:1_FA16:0) | TG | * **↓** | n.s. | n.s. | ** **↓** | *** **↑** | *** **↑** |
| TG(52:1_FA16:1) | TG | n.s. | n.s. | n.s. | ** **↓** | *** **↑** | *** **↑** |
| TG(52:1_FA18:0) | TG | ** **↓** | n.s. | n.s. | ** **↓** | *** **↑** | *** **↑** |
| TG(52:1_FA18:1) | TG | * **↓** | n.s. | n.s. | ** **↓** | *** **↑** | *** **↑** |
| TG(52:1_FA20:0) | TG | n.s. | n.s. | n.s. | * **↓** | *** **↑** | *** **↑** |
| TG(52:1_FA20:1) | TG | ** **↓** | n.s. | n.s. | * **↓** | *** **↑** | *** **↑** |
| TG(52:2_FA14:0) | TG | * **↓** | n.s. | n.s. | *** **↓** | *** **↑** | *** **↑** |
| TG(52:2_FA16:0) | TG | n.s. | n.s. | n.s. | * **↓** | *** **↑** | *** **↑** |
| TG(52:2_FA16:1) | TG | n.s. | n.s. | n.s. | ** **↓** | *** **↑** | *** **↑** |
| TG(52:2_FA18:0) | TG | ** **↓** | n.s. | n.s. | ** **↓** | *** **↑** | *** **↑** |
| TG(52:2_FA18:1) | TG | n.s. | n.s. | n.s. | n.s. | *** **↑** | n.s. |
| TG(52:2_FA18:2) | TG | ** **↓** | n.s. | n.s. | ** **↓** | *** **↑** | *** **↑** |
| TG(52:2_FA20:0) | TG | ** **↓** | n.s. | n.s. | * **↓** | *** **↑** | *** **↑** |
| TG(52:2_FA20:1) | TG | * **↓** | n.s. | n.s. | ** **↓** | *** **↑** | *** **↑** |
| TG(52:2_FA20:2) | TG | * **↓** | n.s. | n.s. | * **↓** | *** **↑** | *** **↑** |
| TG(52:3_FA14:0) | TG | * **↓** | n.s. | n.s. | *** **↓** | *** **↑** | *** **↑** |
| TG(52:3_FA16:0) | TG | n.s. | n.s. | n.s. | * **↓** | *** **↑** | *** **↑** |
| TG(52:3_FA16:1) | TG | n.s. | n.s. | n.s. | n.s. | *** **↑** | ** **↑** |
| TG(52:3_FA18:0) | TG | * **↓** | n.s. | n.s. | ** **↓** | *** **↑** | *** **↑** |
| TG(52:3_FA18:1) | TG | n.s. | n.s. | n.s. | * **↓** | *** **↑** | * **↑** |
| TG(52:3_FA18:2) | TG | n.s. | n.s. | n.s. | * **↓** | *** **↑** | *** **↑** |
| TG(52:3_FA18:3) | TG | * **↓** | n.s. | n.s. | * **↓** | *** **↑** | *** **↑** |
| TG(52:3_FA20:0) | TG | n.s. | n.s. | n.s. | ** **↓** | *** **↑** | *** **↑** |
| TG(52:3_FA20:1) | TG | * **↓** | n.s. | n.s. | ** **↓** | *** **↑** | *** **↑** |
| TG(52:3_FA20:2) | TG | n.s. | n.s. | n.s. | ** **↓** | *** **↑** | *** **↑** |
| TG(52:3_FA20:3) | TG | n.s. | n.s. | n.s. | * **↓** | *** **↑** | *** **↑** |
| TG(52:4_FA14:0) | TG | * **↓** | n.s. | n.s. | * **↓** | *** **↑** | *** **↑** |
| TG(52:4_FA16:0) | TG | * **↓** | n.s. | n.s. | ** **↓** | *** **↑** | *** **↑** |
| TG(52:4_FA16:1) | TG | ** **↓** | n.s. | n.s. | ** **↓** | *** **↑** | *** **↑** |
| TG(52:4_FA18:0) | TG | ** **↓** | n.s. | n.s. | ** **↓** | *** **↑** | *** **↑** |
| TG(52:4_FA18:1) | TG | * **↓** | n.s. | n.s. | ** **↓** | *** **↑** | *** **↑** |
| TG(52:4_FA18:2) | TG | * **↓** | n.s. | n.s. | ** **↓** | *** **↑** | *** **↑** |
| TG(52:4_FA18:3) | TG | * **↓** | n.s. | n.s. | ** **↓** | *** **↑** | *** **↑** |
| TG(52:4_FA20:0) | TG | * **↓** | n.s. | n.s. | ** **↓** | *** **↑** | *** **↑** |
| TG(52:4_FA20:2) | TG | * **↓** | n.s. | n.s. | ** **↓** | *** **↑** | *** **↑** |
| TG(52:4_FA20:3) | TG | * **↓** | n.s. | n.s. | * **↓** | *** **↑** | *** **↑** |
| TG(52:4_FA20:4) | TG | n.s. | n.s. | n.s. | n.s. | *** **↑** | *** **↑** |
| TG(52:4_FA22:4) | TG | n.s. | n.s. | n.s. | * **↓** | *** **↑** | *** **↑** |
| TG(52:5_FA14:0) | TG | n.s. | n.s. | n.s. | n.s. | *** **↑** | *** **↑** |
| TG(52:5_FA16:0) | TG | * **↓** | n.s. | n.s. | ** **↓** | *** **↑** | *** **↑** |
| TG(52:5_FA16:1) | TG | ** **↓** | n.s. | n.s. | ** **↓** | *** **↑** | *** **↑** |
| TG(52:5_FA18:1) | TG | * **↓** | n.s. | n.s. | ** **↓** | *** **↑** | *** **↑** |
| TG(52:5_FA18:2) | TG | * **↓** | n.s. | n.s. | ** **↓** | *** **↑** | *** **↑** |
| TG(52:5_FA18:3) | TG | * **↓** | n.s. | n.s. | ** **↓** | *** **↑** | *** **↑** |
| TG(52:5_FA20:3) | TG | n.s. | n.s. | n.s. | * **↓** | *** **↑** | *** **↑** |
| TG(52:5_FA20:4) | TG | n.s. | n.s. | n.s. | * **↓** | *** **↑** | *** **↑** |
| TG(52:5_FA20:5) | TG | n.s. | n.s. | n.s. | * **↓** | *** **↑** | *** **↑** |
| TG(52:5_FA22:5) | TG | n.s. | n.s. | n.s. | ** **↓** | *** **↑** | *** **↑** |
| TG(52:6_FA14:0) | TG | * **↓** | n.s. | n.s. | * **↓** | *** **↑** | *** **↑** |
| TG(52:6_FA16:0) | TG | * **↓** | n.s. | n.s. | * **↓** | *** **↑** | *** **↑** |
| TG(52:6_FA16:1) | TG | * **↓** | n.s. | n.s. | * **↓** | *** **↑** | *** **↑** |
| TG(52:6_FA18:1) | TG | * **↓** | n.s. | n.s. | * **↓** | *** **↑** | *** **↑** |
| TG(52:6_FA18:2) | TG | * **↓** | n.s. | n.s. | ** **↓** | *** **↑** | *** **↑** |
| TG(52:6_FA18:3) | TG | * **↓** | n.s. | n.s. | ** **↓** | *** **↑** | *** **↑** |
| TG(52:6_FA20:4) | TG | n.s. | n.s. | n.s. | n.s. | *** **↑** | *** **↑** |
| TG(52:6_FA20:5) | TG | n.s. | n.s. | n.s. | n.s. | *** **↑** | *** **↑** |
| TG(52:6_FA22:6) | TG | n.s. | n.s. | n.s. | * **↓** | *** **↑** | *** **↑** |
| TG(52:7_FA16:0) | TG | n.s. | n.s. | n.s. | n.s. | *** **↑** | *** **↑** |
| TG(52:7_FA18:1) | TG | * **↓** | n.s. | n.s. | * **↓** | *** **↑** | *** **↑** |
| TG(52:7_FA20:5) | TG | n.s. | n.s. | n.s. | * **↓** | *** **↑** | *** **↑** |
| TG(52:7_FA22:6) | TG | n.s. | n.s. | n.s. | * **↓** | *** **↑** | *** **↑** |
| TG(52:8_FA16:1) | TG | n.s. | n.s. | n.s. | n.s. | *** **↑** | *** **↑** |
| TG(52:8_FA18:2) | TG | n.s. | n.s. | n.s. | * **↓** | *** **↑** | *** **↑** |
| TG(53:0_FA16:0) | TG | n.s. | n.s. | n.s. | n.s. | *** **↑** | *** **↑** |
| TG(53:1_FA16:0) | TG | n.s. | n.s. | n.s. | ***↓** | *** **↑** | *** **↑** |
| TG(53:1_FA17:0) | TG | n.s. | n.s. | n.s. | ** **↓** | *** **↑** | *** **↑** |
| TG(53:1_FA18:0) | TG | ***↓** | n.s. | n.s. | ** **↓** | *** **↑** | *** **↑** |
| TG(53:1_FA18:1) | TG | n.s. | n.s. | n.s. | * **↓** | *** **↑** | *** **↑** |
| TG(53:2_FA16:0) | TG | n.s. | n.s. | n.s. | ** **↓** | *** **↑** | *** **↑** |
| TG(53:2_FA17:0) | TG | * **↓** | n.s. | n.s. | ** **↓** | *** **↑** | *** **↑** |
| TG(53:2_FA18:1) | TG | * **↓** | n.s. | n.s. | ** **↓** | *** **↑** | *** **↑** |
| TG(53:2_FA18:2) | TG | * **↓** | n.s. | n.s. | ** **↓** | *** **↑** | *** **↑** |
| TG(53:3_FA16:0) | TG | n.s. | n.s. | n.s. | * **↓** | *** **↑** | *** **↑** |
| TG(53:3_FA17:0) | TG | n.s. | n.s. | n.s. | * **↓** | *** **↑** | *** **↑** |
| TG(53:3_FA18:2) | TG | * **↓** | n.s. | n.s. | ** **↓** | *** **↑** | *** **↑** |
| TG(53:4_FA16:0) | TG | * **↓** | n.s. | n.s. | ** **↓** | *** **↑** | *** **↑** |
| TG(53:4_FA17:0) | TG | * **↓** | n.s. | n.s. | ** **↓** | *** **↑** | *** **↑** |
| TG(53:4_FA18:2) | TG | * **↓** | n.s. | n.s. | ** **↓** | *** **↑** | *** **↑** |
| TG(53:4_FA18:3) | TG | * **↓** | n.s. | n.s. | ** **↓** | *** **↑** | *** **↑** |
| TG(53:4_FA20:4) | TG | n.s. | n.s. | n.s. | n.s. | *** **↑** | *** **↑** |
| TG(53:5_FA20:4) | TG | n.s. | n.s. | n.s. | n.s. | *** **↑** | *** **↑** |
| TG(53:6_FA20:4) | TG | n.s. | n.s. | n.s. | n.s. | *** **↑** | *** **↑** |
| TG(54:0_FA16:0) | TG | n.s. | n.s. | n.s. | * **↓** | *** **↑** | *** **↑** |
| TG(54:0_FA18:0) | TG | * **↓** | n.s. | n.s. | ** **↓** | *** **↑** | *** **↑** |
| TG(54:1_FA16:0) | TG | n.s. | n.s. | n.s. | * **↓** | *** **↑** | *** **↑** |
| TG(54:1_FA18:0) | TG | * **↓** | n.s. | n.s. | ** **↓** | *** **↑** | *** **↑** |
| TG(54:1_FA18:1) | TG | n.s. | n.s. | n.s. | ** **↓** | *** **↑** | *** **↑** |
| TG(54:1_FA20:0) | TG | n.s. | n.s. | n.s. | * **↓** | *** **↑** | *** **↑** |
| TG(54:1_FA20:1) | TG | * **↓** | n.s. | n.s. | ** **↓** | *** **↑** | *** **↑** |
| TG(54:2_FA16:0) | TG | * **↓** | n.s. | n.s. | ** **↓** | *** **↑** | *** **↑** |
| TG(54:2_FA18:0) | TG | n.s. | n.s. | n.s. | ** **↓** | *** **↑** | *** **↑** |
| TG(54:2_FA18:1) | TG | n.s. | n.s. | n.s. | ** **↓** | *** **↑** | *** **↑** |
| TG(54:2_FA18:2) | TG | ** **↓** | n.s. | n.s. | ** **↓** | *** **↑** | *** **↑** |
| TG(54:2_FA20:0) | TG | n.s. | n.s. | n.s. | * **↓** | *** **↑** | *** **↑** |
| TG(54:2_FA20:1) | TG | * **↓** | n.s. | n.s. | ** **↓** | *** **↑** | *** **↑** |
| TG(54:2_FA20:2) | TG | * **↓** | n.s. | n.s. | * **↓** | *** **↑** | *** **↑** |
| TG(54:3_FA16:0) | TG | * **↓** | n.s. | n.s. | ** **↓** | *** **↑** | *** **↑** |
| TG(54:3_FA16:1) | TG | * **↓** | n.s. | n.s. | ** **↓** | *** **↑** | ** **↑** |
| TG(54:3_FA18:0) | TG | * **↓** | n.s. | n.s. | ** **↓** | *** **↑** | *** **↑** |
| TG(54:3_FA18:1) | TG | n.s. | n.s. | n.s. | ** **↓** | *** **↑** | ** **↑** |
| TG(54:3_FA18:2) | TG | * **↓** | n.s. | n.s. | ** **↓** | *** **↑** | *** **↑** |
| TG(54:3_FA18:3) | TG | ** **↓** | n.s. | n.s. | ** **↓** | *** **↑** | *** **↑** |
| TG(54:3_FA20:1) | TG | * **↓** | n.s. | n.s. | ** **↓** | *** **↑** | *** **↑** |
| TG(54:3_FA20:2) | TG | n.s. | n.s. | n.s. | ** **↓** | *** **↑** | *** **↑** |
| TG(54:3_FA20:3) | TG | n.s. | n.s. | n.s. | * **↓** | *** **↑** | *** **↑** |
| TG(54:4_FA16:0) | TG | * **↓** | n.s. | n.s. | ** **↓** | *** **↑** | *** **↑** |
| TG(54:4_FA16:1) | TG | * **↓** | n.s. | n.s. | ** **↓** | *** **↑** | *** **↑** |
| TG(54:4_FA18:0) | TG | * **↓** | n.s. | n.s. | ** **↓** | *** **↑** | *** **↑** |
| TG(54:4_FA18:1) | TG | n.s. | n.s. | n.s. | ** **↓** | *** **↑** | *** **↑** |
| TG(54:4_FA18:2) | TG | n.s. | n.s. | n.s. | ** **↓** | *** **↑** | *** **↑** |
| TG(54:4_FA18:3) | TG | * **↓** | n.s. | n.s. | * **↓** | *** **↑** | *** **↑** |
| TG(54:4_FA20:1) | TG | * **↓** | n.s. | n.s. | ** **↓** | *** **↑** | *** **↑** |
| TG(54:4_FA20:2) | TG | * **↓** | n.s. | n.s. | ** **↓** | *** **↑** | *** **↑** |
| TG(54:4_FA20:3) | TG | n.s. | n.s. | n.s. | * **↓** | *** **↑** | *** **↑** |
| TG(54:4_FA20:4) | TG | n.s. | n.s. | n.s. | * **↓** | *** **↑** | *** **↑** |
| TG(54:4_FA22:4) | TG | n.s. | n.s. | n.s. | n.s. | *** **↑** | *** **↑** |
| TG(54:5_FA16:0) | TG | * **↓** | n.s. | n.s. | ** **↓** | *** **↑** | *** **↑** |
| TG(54:5_FA16:1) | TG | * **↓** | n.s. | n.s. | n.s. | *** **↑** | *** **↑** |
| TG(54:5_FA18:0) | TG | ** **↓** | n.s. | n.s. | ** **↓** | *** **↑** | *** **↑** |
| TG(54:5_FA18:1) | TG | * **↓** | n.s. | n.s. | ** **↓** | *** **↑** | *** **↑** |
| TG(54:5_FA18:2) | TG | * **↓** | n.s. | n.s. | ** **↓** | *** **↑** | *** **↑** |
| TG(54:5_FA18:3) | TG | * **↓** | n.s. | n.s. | ** **↓** | *** **↑** | *** **↑** |
| TG(54:5_FA20:2) | TG | ** **↓** | n.s. | n.s. | ** **↓** | *** **↑** | *** **↑** |
| TG(54:5_FA20:3) | TG | n.s. | n.s. | n.s. | * **↓** | *** **↑** | *** **↑** |
| TG(54:5_FA20:4) | TG | n.s. | n.s. | n.s. | ** **↓** | *** **↑** | *** **↑** |
| TG(54:5_FA20:5) | TG | n.s. | n.s. | n.s. | n.s. | *** **↑** | *** **↑** |
| TG(54:5_FA22:4) | TG | * **↓** | n.s. | n.s. | * **↓** | *** **↑** | *** **↑** |
| TG(54:5_FA22:5) | TG | * **↓** | n.s. | n.s. | * **↓** | *** **↑** | *** **↑** |
| TG(54:6_FA16:0) | TG | n.s. | n.s. | n.s. | * **↓** | *** **↑** | *** **↑** |
| TG(54:6_FA16:1) | TG | n.s. | n.s. | n.s. | * **↓** | *** **↑** | *** **↑** |
| TG(54:6_FA18:1) | TG | * **↓** | n.s. | n.s. | ** **↓** | *** **↑** | *** **↑** |
| TG(54:6_FA18:2) | TG | * **↓** | n.s. | n.s. | ** **↓** | *** **↑** | *** **↑** |
| TG(54:6_FA18:3) | TG | * **↓** | n.s. | n.s. | ** **↓** | *** **↑** | *** **↑** |
| TG(54:6_FA20:3) | TG | * **↓** | n.s. | n.s. | * **↓** | *** **↑** | *** **↑** |
| TG(54:6_FA20:4) | TG | n.s. | n.s. | n.s. | * **↓** | *** **↑** | *** **↑** |
| TG(54:6_FA20:5) | TG | n.s. | n.s. | n.s. | n.s. | *** **↑** | *** **↑** |
| TG(54:6_FA22:5) | TG | * **↓** | n.s. | n.s. | n.s. | *** **↑** | *** **↑** |
| TG(54:6_FA22:6) | TG | * **↓** | n.s. | n.s. | * **↓** | *** **↑** | *** **↑** |
| TG(54:7_FA16:1) | TG | n.s. | n.s. | n.s. | n.s. | *** **↑** | *** **↑** |
| TG(54:7_FA18:1) | TG | * **↓** | n.s. | n.s. | * **↓** | *** **↑** | *** **↑** |
| TG(54:7_FA18:2) | TG | * **↓** | n.s. | n.s. | ** **↓** | *** **↑** | *** **↑** |
| TG(54:7_FA18:3) | TG | * **↓** | n.s. | n.s. | ** **↓** | *** **↑** | *** **↑** |
| TG(54:7_FA20:4) | TG | n.s. | n.s. | n.s. | n.s. | *** **↑** | *** **↑** |
| TG(54:7_FA20:5) | TG | n.s. | n.s. | n.s. | n.s. | *** **↑** | *** **↑** |
| TG(54:7_FA22:5) | TG | n.s. | n.s. | n.s. | * **↓** | *** **↑** | *** **↑** |
| TG(54:7_FA22:6) | TG | n.s. | n.s. | n.s. | * **↓** | *** **↑** | *** **↑** |
| TG(54:8_FA18:2) | TG | n.s. | n.s. | n.s. | ** **↓** | *** **↑** | *** **↑** |
| TG(54:8_FA18:3) | TG | * **↓** | n.s. | n.s. | * **↓** | *** **↑** | *** **↑** |
| TG(54:8_FA20:4) | TG | n.s. | n.s. | n.s. | n.s. | *** **↑** | *** **↑** |
| TG(54:8_FA20:5) | TG | n.s. | n.s. | n.s. | n.s. | *** **↑** | *** **↑** |
| TG(54:8_FA22:6) | TG | n.s. | n.s. | n.s. | * **↓** | *** **↑** | *** **↑** |
| TG(55:1_FA16:0) | TG | n.s. | n.s. | n.s. | n.s. | *** **↑** | *** **↑** |
| TG(55:1_FA18:1) | TG | * **↓** | n.s. | n.s. | * **↓** | *** **↑** | *** **↑** |
| TG(55:2_FA18:1) | TG | n.s. | n.s. | n.s. | ** **↓** | *** **↑** | *** **↑** |
| TG(55:2_FA18:2) | TG | n.s. | n.s. | n.s. | ** **↓** | *** **↑** | *** **↑** |
| TG(55:3_FA18:1) | TG | n.s. | n.s. | n.s. | ** **↓** | *** **↑** | *** **↑** |
| TG(55:3_FA18:2) | TG | n.s. | n.s. | n.s. | ** **↓** | *** **↑** | *** **↑** |
| TG(55:4_FA18:1) | TG | * **↓** | n.s. | n.s. | ** **↓** | *** **↑** | *** **↑** |
| TG(55:4_FA18:2) | TG | n.s. | n.s. | n.s. | ** **↓** | *** **↑** | *** **↑** |
| TG(55:5_FA18:1) | TG | n.s. | n.s. | n.s. | * **↓** | *** **↑** | *** **↑** |
| TG(55:5_FA20:4) | TG | n.s. | n.s. | n.s. | * **↓** | *** **↑** | *** **↑** |
| TG(55:5_FA22:6) | TG | * **↓** | n.s. | n.s. | ** **↓** | *** **↑** | *** **↑** |
| TG(56:1_FA16:0) | TG | * **↓** | n.s. | n.s. | n.s. | *** **↑** | *** **↑** |
| TG(56:1_FA18:1) | TG | * **↓** | n.s. | n.s. | * **↓** | *** **↑** | *** **↑** |
| TG(56:10_FA18:2) | TG | n.s. | n.s. | n.s. | ** **↓** | *** **↑** | *** **↑** |
| TG(56:2_FA16:0) | TG | * **↓** | n.s. | n.s. | * **↓** | *** **↑** | *** **↑** |
| TG(56:2_FA18:0) | TG | * **↓** | n.s. | n.s. | ** **↓** | *** **↑** | *** **↑** |
| TG(56:2_FA20:0) | TG | n.s. | n.s. | n.s. | * **↓** | *** **↑** | *** **↑** |
| TG(56:2_FA20:1) | TG | * **↓** | n.s. | n.s. | ** **↓** | *** **↑** | *** **↑** |
| TG(56:3_FA16:0) | TG | n.s. | n.s. | n.s. | ** **↓** | *** **↑** | *** **↑** |
| TG(56:3_FA18:0) | TG | * **↓** | n.s. | n.s. | ** **↓** | *** **↑** | *** **↑** |
| TG(56:3_FA18:1) | TG | * **↓** | n.s. | n.s. | ** **↓** | *** **↑** | *** **↑** |
| TG(56:3_FA18:2) | TG | n.s. | n.s. | n.s. | ** **↓** | *** **↑** | *** **↑** |
| TG(56:3_FA20:0) | TG | n.s. | n.s. | n.s. | * **↓** | *** **↑** | *** **↑** |
| TG(56:3_FA20:1) | TG | n.s. | n.s. | n.s. | ** **↓** | *** **↑** | *** **↑** |
| TG(56:3_FA20:2) | TG | * **↓** | n.s. | n.s. | ** **↓** | *** **↑** | ** **↑** |
| TG(56:4_FA16:0) | TG | * **↓** | n.s. | n.s. | * **↓** | *** **↑** | *** **↑** |
| TG(56:4_FA18:0) | TG | * **↓** | n.s. | n.s. | * **↓** | *** **↑** | *** **↑** |
| TG(56:4_FA18:1) | TG | * **↓** | n.s. | n.s. | ** **↓** | *** **↑** | ** **↑** |
| TG(56:4_FA18:2) | TG | * **↓** | n.s. | n.s. | ** **↓** | *** **↑** | *** **↑** |
| TG(56:4_FA20:1) | TG | * **↓** | n.s. | n.s. | ** **↓** | *** **↑** | *** **↑** |
| TG(56:4_FA20:2) | TG | * **↓** | n.s. | n.s. | ** **↓** | *** **↑** | ** **↑** |
| TG(56:4_FA20:3) | TG | * **↓** | n.s. | n.s. | * **↓** | *** **↑** | *** **↑** |
| TG(56:4_FA20:4) | TG | * **↓** | n.s. | n.s. | ** **↓** | *** **↑** | ** **↑** |
| TG(56:4_FA22:4) | TG | * **↓** | n.s. | n.s. | n.s. | *** **↑** | *** **↑** |
| TG(56:5_FA16:0) | TG | n.s. | n.s. | n.s. | * **↓** | *** **↑** | *** **↑** |
| TG(56:5_FA18:0) | TG | * **↓** | n.s. | n.s. | ** **↓** | *** **↑** | ** **↑** |
| TG(56:5_FA18:1) | TG | * **↓** | n.s. | n.s. | ** **↓** | *** **↑** | *** **↑** |
| TG(56:5_FA18:2) | TG | * **↓** | n.s. | n.s. | ** **↓** | *** **↑** | *** **↑** |
| TG(56:5_FA20:1) | TG | * **↓** | n.s. | n.s. | ** **↓** | *** **↑** | *** **↑** |
| TG(56:5_FA20:2) | TG | * **↓** | n.s. | n.s. | ** **↓** | *** **↑** | *** **↑** |
| TG(56:5_FA20:3) | TG | n.s. | n.s. | n.s. | * **↓** | *** **↑** | *** **↑** |
| TG(56:5_FA20:4) | TG | n.s. | n.s. | n.s. | *** **↓** | *** **↑** | n.s. |
| TG(56:5_FA22:4) | TG | n.s. | n.s. | n.s. | * **↓** | *** **↑** | *** **↑** |
| TG(56:5_FA22:5) | TG | * **↓** | n.s. | n.s. | *** **↓** | *** **↑** | ** **↑** |
| TG(56:6_FA16:0) | TG | * **↓** | n.s. | n.s. | ** **↓** | *** **↑** | ** **↑** |
| TG(56:6_FA18:1) | TG | * **↓** | n.s. | n.s. | ** **↓** | *** **↑** | ** **↑** |
| TG(56:6_FA18:2) | TG | * **↓** | n.s. | n.s. | ** **↓** | *** **↑** | *** **↑** |
| TG(56:6_FA18:3) | TG | ** **↓** | n.s. | n.s. | * **↓** | *** **↑** | *** **↑** |
| TG(56:6_FA20:2) | TG | n.s. | n.s. | n.s. | ** **↓** | *** **↑** | *** **↑** |
| TG(56:6_FA20:3) | TG | n.s. | n.s. | n.s. | * **↓** | *** **↑** | ** **↑** |
| TG(56:6_FA20:4) | TG | n.s. | n.s. | n.s. | * **↓** | *** **↑** | * **↑** |
| TG(56:6_FA20:5) | TG | * **↓** | n.s. | n.s. | n.s. | * **↑** | n.s. |
| TG(56:6_FA22:4) | TG | n.s. | n.s. | n.s. | * **↓** | *** **↑** | *** **↑** |
| TG(56:6_FA22:5) | TG | * **↓** | n.s. | n.s. | ** **↓** | *** **↑** | ** **↑** |
| TG(56:6_FA22:6) | TG | n.s. | n.s. | n.s. | ** **↓** | *** **↑** | ** **↑** |
| TG(56:7_FA16:0) | TG | n.s. | n.s. | n.s. | n.s. | *** **↑** | ** **↑** |
| TG(56:7_FA16:1) | TG | * **↓** | n.s. | n.s. | * **↓** | *** **↑** | *** **↑** |
| TG(56:7_FA18:0) | TG | n.s. | n.s. | n.s. | * **↓** | *** **↑** | ** **↑** |
| TG(56:7_FA18:1) | TG | n.s. | n.s. | n.s. | n.s. | ** **↑** | n.s. |
| TG(56:7_FA18:2) | TG | n.s. | n.s. | n.s. | * **↓** | *** **↑** | *** **↑** |
| TG(56:7_FA18:3) | TG | * **↓** | n.s. | n.s. | n.s. | *** **↑** | ** **↑** |
| TG(56:7_FA20:3) | TG | n.s. | n.s. | n.s. | n.s. | *** **↑** | *** **↑** |
| TG(56:7_FA20:4) | TG | n.s. | n.s. | n.s. | n.s. | *** **↑** | * **↑** |
| TG(56:7_FA20:5) | TG | n.s. | n.s. | n.s. | n.s. | ** **↑** | n.s. |
| TG(56:7_FA22:4) | TG | * **↓** | n.s. | n.s. | * **↓** | *** **↑** | *** **↑** |
| TG(56:7_FA22:5) | TG | n.s. | n.s. | n.s. | n.s. | *** **↑** | *** **↑** |
| TG(56:7_FA22:6) | TG | n.s. | n.s. | n.s. | * **↓** | *** **↑** | ** **↑** |
| TG(56:8_FA16:0) | TG | * **↓** | n.s. | n.s. | * **↓** | *** **↑** | *** **↑** |
| TG(56:8_FA16:1) | TG | n.s. | n.s. | n.s. | n.s. | *** **↑** | ** **↑** |
| TG(56:8_FA18:1) | TG | n.s. | n.s. | n.s. | n.s. | *** **↑** | ** **↑** |
| TG(56:8_FA18:2) | TG | n.s. | n.s. | n.s. | * **↓** | *** **↑** | ** **↑** |
| TG(56:8_FA18:3) | TG | n.s. | n.s. | n.s. | n.s. | *** **↑** | *** **↑** |
| TG(56:8_FA20:4) | TG | n.s. | n.s. | n.s. | n.s. | *** **↑** | *** **↑** |
| TG(56:8_FA22:5) | TG | * **↓** | n.s. | n.s. | ** **↓** | *** **↑** | *** **↑** |
| TG(56:8_FA22:6) | TG | n.s. | n.s. | n.s. | * **↓** | *** **↑** | *** **↑** |
| TG(56:9_FA18:3) | TG | n.s. | n.s. | n.s. | n.s. | *** **↑** | *** **↑** |
| TG(56:9_FA20:4) | TG | n.s. | n.s. | n.s. | n.s. | *** **↑** | *** **↑** |
| TG(56:9_FA20:5) | TG | n.s. | n.s. | n.s. | n.s. | *** **↑** | *** **↑** |
| TG(56:9_FA22:6) | TG | n.s. | n.s. | n.s. | * **↓** | *** **↑** | *** **↑** |
| TG(57:2_FA18:1) | TG | n.s. | n.s. | n.s. | * **↓** | *** **↑** | *** **↑** |
| TG(57:2_FA18:2) | TG | n.s. | n.s. | n.s. | * **↓** | *** **↑** | *** **↑** |
| TG(57:3_FA18:2) | TG | n.s. | n.s. | n.s. | * **↓** | *** **↑** | *** **↑** |
| TG(58:10_FA18:2) | TG | n.s. | n.s. | n.s. | n.s. | *** **↑** | ** **↑** |
| TG(58:10_FA20:5) | TG | n.s. | n.s. | n.s. | n.s. | *** **↑** | * **↑** |
| TG(58:10_FA22:5) | TG | n.s. | n.s. | n.s. | n.s. | *** **↑** | *** **↑** |
| TG(58:10_FA22:6) | TG | n.s. | n.s. | n.s. | n.s. | *** **↑** | *** **↑** |
| TG(58:2_FA18:1) | TG | n.s. | n.s. | n.s. | * **↓** | *** **↑** | *** **↑** |
| TG(58:3_FA18:1) | TG | n.s. | n.s. | n.s. | * **↓** | *** **↑** | *** **↑** |
| TG(58:5_FA18:1) | TG | n.s. | n.s. | n.s. | * **↓** | *** **↑** | *** **↑** |
| TG(58:6_FA16:0) | TG | n.s. | n.s. | n.s. | * **↓** | *** **↑** | ** **↑** |
| TG(58:6_FA18:0) | TG | ** **↓** | n.s. | n.s. | ** **↓** | *** **↑** | ** **↑** |
| TG(58:6_FA18:1) | TG | n.s. | n.s. | n.s. | * **↓** | *** **↑** | *** **↑** |
| TG(58:6_FA20:4) | TG | n.s. | n.s. | n.s. | n.s. | *** **↑** | n.s. |
| TG(58:6_FA22:4) | TG | n.s. | n.s. | n.s. | n.s. | *** **↑** | *** **↑** |
| TG(58:6_FA22:5) | TG | * **↓** | n.s. | n.s. | ** **↓** | *** **↑** | ** **↑** |
| TG(58:7_FA16:0) | TG | n.s. | n.s. | n.s. | * **↓** | *** **↑** | ** **↑** |
| TG(58:7_FA18:0) | TG | n.s. | n.s. | n.s. | * **↓** | *** **↑** | ** **↑** |
| TG(58:7_FA18:1) | TG | n.s. | n.s. | n.s. | * **↓** | *** **↑** | ** **↑** |
| TG(58:7_FA18:2) | TG | * **↓** | n.s. | n.s. | n.s. | *** **↑** | *** **↑** |
| TG(58:7_FA20:4) | TG | n.s. | n.s. | n.s. | n.s. | * **↑** | n.s. |
| TG(58:7_FA22:4) | TG | n.s. | n.s. | n.s. | * **↓** | *** **↑** | ** **↑** |
| TG(58:7_FA22:5) | TG | n.s. | n.s. | n.s. | * **↓** | *** **↑** | ** **↑** |
| TG(58:7_FA22:6) | TG | n.s. | n.s. | n.s. | ** **↓** | *** **↑** | ** **↑** |
| TG(58:8_FA18:1) | TG | n.s. | n.s. | n.s. | n.s. | ** **↑** | * **↑** |
| TG(58:8_FA18:2) | TG | n.s. | n.s. | n.s. | n.s. | *** **↑** | * **↑** |
| TG(58:8_FA20:3) | TG | n.s. | n.s. | n.s. | n.s. | *** **↑** | n.s. |
| TG(58:8_FA20:4) | TG | n.s. | n.s. | n.s. | n.s. | ** **↑** | ** **↑** |
| TG(58:8_FA22:5) | TG | n.s. | n.s. | n.s. | n.s. | *** **↑** | n.s. |
| TG(58:8_FA22:6) | TG | n.s. | n.s. | n.s. | n.s. | *** **↑** | * **↑** |
| TG(58:9_FA18:1) | TG | n.s. | n.s. | n.s. | n.s. | *** **↑** | ** **↑** |
| TG(58:9_FA18:2) | TG | n.s. | n.s. | n.s. | n.s. | *** **↑** | ** **↑** |
| TG(58:9_FA20:4) | TG | n.s. | n.s. | n.s. | n.s. | ** **↑** | ** **↑** |
| TG(58:9_FA22:5) | TG | n.s. | n.s. | n.s. | n.s. | *** **↑** | *** **↑** |
| TG(58:9_FA22:6) | TG | n.s. | n.s. | n.s. | n.s. | *** **↑** | ** **↑** |
| TG(60:10_FA22:5) | TG | n.s. | n.s. | n.s. | n.s. | *** **↑** | * **↑** |
| TG(60:11_FA22:5) | TG | n.s. | n.s. | n.s. | n.s. | ** **↑** | * **↑** |
| TG(60:11_FA22:5) | TG | n.s. | n.s. | n.s. | n.s. | *** **↑** | * **↑** |

**Table S5 Significant lipoproteins with an adjusted *p*-value < 0.05 from pairwise Wilcoxon signed-rank test.** Significance denoted by * = *p* < 0.05, ** = *p* < 0.01, *** = *p* < 0.001.

|  | Sampling site comparison adjusted *p*-value | | | | | |
| --- | --- | --- | --- | --- | --- | --- |
| Lipoprotein | **A1 ~ A2** | **A1 ~ CS** | **A2 ~ RRV** | **CS ~ RRV** | **RRV ~ PV** | **CS ~ PV** |
| TPTG | * **↓** | * **↓** | n.s. | * **↓** | *** **↑** | *** **↑** |
| TPCH | * **↑** | n.s. | n.s. | n.s. | * **↑** | ** **↑** |
| LDCH | * **↑** | n.s. | n.s. | n.s. | n.s. | * **↑** |
| HDCH | * **↑** | n.s. | n.s. | n.s. | n.s. | n.s. |
| TPA2 | ** **↑** | n.s. | n.s. | * **↑** | ** **↓** | n.s. |
| TPAB | n.s. | n.s. | n.s. | n.s. | *** **↑** | *** **↑** |
| ABA1 | n.s. | n.s. | * **↑** | n.s. | *** **↑** | *** **↑** |
| TBPN | n.s. | n.s. | n.s. | n.s. | *** **↑** | *** **↑** |
| VLPN | * **↓** | * **↓** | n.s. | n.s. | *** **↑** | *** **↑** |
| IDPN | n.s. | n.s. | n.s. | n.s. | *** **↑** | *** **↑** |
| LDPN | n.s. | n.s. | n.s. | n.s. | *** **↑** | *** **↑** |
| L2PN | ** **↑** | n.s. | n.s. | * **↑** | *** **↓** | n.s. |
| L3PN | ** **↑** | * **↑** | n.s. | n.s. | ** **↓** | n.s. |
| L5PN | n.s. | n.s. | n.s. | n.s. | *** **↑** | *** **↑** |
| L6PN | n.s. | n.s. | n.s. | n.s. | *** **↑** | *** **↑** |
| VLTG | * **↓** | n.s. | n.s. | n.s. | *** **↑** | *** **↑** |
| IDTG | * **↓** | n.s. | n.s. | * **↓** | *** **↑** | *** **↑** |
| LDTG | n.s. | n.s. | n.s. | n.s. | *** **↑** | *****↑** |
| HDTG | n.s. | * **↓** | * **↓** | n.s. | ** **↑** | ** **↑** |
| VLCH | n.s. | n.s. | n.s. | n.s. | *** **↑** | *** **↑** |
| IDCH | n.s. | n.s. | n.s. | n.s. | *** **↑** | *** **↑** |
| VLFC | * **↓** | * **↓** | n.s. | n.s. | *** **↑** | *** **↑** |
| IDFC | n.s. | n.s. | n.s. | n.s. | *** **↑** | ** **↑** |
| LDFC | * **↑** | n.s. | n.s. | * **↑** | n.s. | n.s. |
| HDFC | ** **↑** | n.s. | n.s. | n.s. | *** **↓** | ***↓** |
| VLPL | * **↓** | n.s. | n.s. | n.s. | *** **↑** | *** **↑** |
| IDPL | n.s. | n.s. | n.s. | n.s. | *** **↑** | *** **↑** |
| LDPL | * **↑** | n.s. | n.s. | * **↑** | n.s. | n.s. |
| HDPL | ** **↑** | n.s. | n.s. | * **↑** | ** **↓** | n.s. |
| HDA1 | * **↑** | n.s. | n.s. | n.s. | n.s. | n.s. |
| HDA2 | ** **↑** | n.s. | n.s. | * **↑** | * **↓** | n.s. |
| VLAB | * **↓** | * **↓** | n.s. | n.s. | *** **↑** | *** **↑** |
| IDAB | n.s. | n.s. | n.s. | n.s. | *** **↑** | *** **↑** |
| LDAB | n.s. | n.s. | n.s. | n.s. | *** **↑** | *** **↑** |
| V1TG | * **↓** | n.s. | n.s. | n.s. | *** **↑** | *** **↑** |
| V2TG | * **↓** | n.s. | n.s. | n.s. | *** **↑** | *** **↑** |
| V3TG | n.s. | n.s. | n.s. | n.s. | *** **↑** | *** **↑** |
| V4TG | n.s. | n.s. | n.s. | n.s. | *** **↑** | *** **↑** |
| V1CH | ** **↓** | n.s. | n.s. | n.s. | *** **↑** | *** **↑** |
| V2CH | n.s. | n.s. | n.s. | n.s. | *** **↑** | *** **↑** |
| V4CH | n.s. | n.s. | n.s. | n.s. | n.s. | * **↑** |
| V5CH | * **↓** | n.s. | n.s. | n.s. | n.s. | n.s. |
| V1FC | * **↓** | n.s. | n.s. | * **↓** | *** **↑** | *** **↑** |
| V2FC | n.s. | n.s. | n.s. | n.s. | *** **↑** | *** **↑** |
| V3FC | n.s. | n.s. | n.s. | n.s. | *** **↑** | *** **↑** |
| V1PL | * **↓** | * **↓** | n.s. | n.s. | *** **↑** | *** **↑** |
| V2PL | n.s. | n.s. | n.s. | n.s. | *** **↑** | *** **↑** |
| V3PL | n.s. | n.s. | n.s. | n.s. | *** **↑** | *** **↑** |
| V4PL | n.s. | n.s. | n.s. | n.s. | * **↑** | ** **↑** |
| V5PL | * **↓** | n.s. | n.s. | n.s. | n.s. | n.s. |
| L1TG | n.s. | n.s. | n.s. | * **↓** | *** **↑** | *** **↑** |
| L2TG | n.s. | *** **↑** | n.s. | n.s. | *** **↑** | *****↑** |
| L3TG | n.s. | n.s. | n.s. | n.s. | * **↑** | * **↑** |
| L4TG | n.s. | n.s. | n.s. | * **↓** | *** **↑** | *** **↑** |
| L5TG | n.s. | n.s. | n.s. | n.s. | *** **↑** | *** **↑** |
| L6TG | n.s. | n.s. | n.s. | n.s. | *** **↑** | *** **↑** |
| L2CH | ** **↑** | * **↑** | n.s. | * **↑** | *** **↓** | n.s. |
| L3CH | ** **↑** | ** **↑** | n.s. | n.s. | *** **↓** | n.s. |
| L5CH | n.s. | n.s. | n.s. | n.s. | *** **↑** | *** **↑** |
| L6CH | n.s. | n.s. | n.s. | n.s. | *** **↑** | *** **↑** |
| L1FC | * **↑** | n.s. | n.s. | n.s. | n.s. | n.s. |
| L2FC | ** **↑** | n.s. | n.s. | * **↑** | ** **↓** | n.s. |
| L3FC | ** **↑** | ** **↑** | n.s. | * | ** **↓** | * **↓** |
| L5FC | n.s. | n.s. | n.s. | n.s. | n.s. | ** **↑** |
| L6FC | n.s. | n.s. | n.s. | n.s. | *** **↑** | *** **↑** |
| L2PL | ** **↑** | * **↑** | n.s. | * **↑** | ** **↓** | * **↓** |
| L3PL | ** **↑** | * **↑** | n.s. | n.s. | *** **↓** | n.s. |
| L5PL | n.s. | n.s. | n.s. | n.s. | ** **↑** | *** **↑** |
| L6PL | n.s. | n.s. | n.s. | n.s. | *** **↑** | *** **↑** |
| L2AB | ** **↑** | n.s. | n.s. | * **↑** | ** **↓** | n.s. |
| L3AB | * **↑** | * **↑** | n.s. | n.s. | ** **↓** | n.s. |
| L5AB | n.s. | n.s. | n.s. | n.s. | *** **↑** | *** **↑** |
| L6AB | n.s. | n.s. | n.s. | n.s. | *** **↑** | *** **↑** |
| H1TG | n.s. | * **↓** | n.s. | n.s. | *** **↑** | *** **↑** |
| H3TG | n.s. | n.s. | * **↓** | n.s. | *** **↑** | ** **↑** |
| H4TG | ** **↓** | * **↓** | n.s. | * **↓** | *** **↑** | *** **↑** |
| H1CH | n.s. | n.s. | n.s. | n.s. | *** **↑** | *** **↑** |
| H2CH | *** **↑** | n.s. | n.s. | ** **↑** | *** **↓** | n.s. |
| H3CH | *** **↑** | n.s. | n.s. | ** **↑** | *** **↓** | * **↓** |
| H4CH | ** **↑** | n.s. | n.s. | * **↑** | ** **↓** | * **↓** |
| H1FC | * **↑** | n.s. | n.s. | n.s. | n.s. | n.s. |
| H2FC | *** **↑** | n.s. | n.s. | * **↑** | n.s. | * **↓** |
| H3FC | ** **↑** | n.s. | n.s. | n.s. | ** **↓** | n.s. |
| H4FC | *** **↑** | n.s. | n.s. | ** **↑** | *** **↓** | ** **↓** |
| H1PL | n.s. | n.s. | n.s. | n.s. | *** **↑** | *** **↑** |
| H2PL | *** **↑** | n.s. | n.s. | * **↑** | *** **↓** | * **↓** |
| H3PL | ** **↑** | n.s. | n.s. | * **↑** | ** **↓** | * **↓** |
| H4PL | n.s. | n.s. | n.s. | n.s. | * **↓** | * **↓** |
| H1A1 | n.s. | n.s. | n.s. | n.s. | *** **↑** | *** **↑** |
| H2A1 | ** **↑** | n.s. | n.s. | * **↑** | * **↓** | n.s. |
| H3A1 | *** **↑** | n.s. | n.s. | ** **↑** | *** **↓** | ** **↓** |
| H4A1 | * **↑** | n.s. | n.s. | * **↑** | * **↓** | n.s. |
| H1A2 | n.s. | n.s. | n.s. | n.s. | * **↑** | * **↑** |
| H3A2 | * **↑** | n.s. | n.s. | n.s. | n.s. | n.s. |
| H4A2 | ** **↑** | n.s. | n.s. | * **↑** | ** **↓** | n.s. |

**Table S6 Significant small molecules with an adjusted *p*-value < 0.05 from pairwise Wilcoxon signed-rank test.** Significance denoted by * = *p* < 0.05, ** = *p* < 0.01, *** = *p* < 0.001.

|  | Sampling site comparison adjusted *p*-value | | | | | |
| --- | --- | --- | --- | --- | --- | --- |
| Small molecule | A1 ~ A2 | A1 ~ CS | A2 ~ RRV | CS ~ RRV | RRV ~ PV | CS ~ PV |
| Alanine | n.s. | n.s. | n.s. | n.s. | *** **↑** | *** **↑** |
| Creatine | n.s. | n.s. | n.s. | n.s. | * **↑** | n.s. |
| Creatinine | n.s. | n.s. | *** **↑** | n.s. | *** **↑** | *** **↑** |
| Glutamic acid | n.s. | *** **↓** | n.s. | n.s. | *** **↓** | *** **↓** |
| Glutamine | n.s. | n.s. | * **↓** | *** **↓** | *** **↑** | *** **↑** |
| Glycine | n.s. | n.s. | n.s. | n.s. | ** **↓** | ** **↓** |
| Methionine | n.s. | n.s. | n.s. | ** **↓** | n.s. | n.s. |
| Acetic acid | n.s. | *** **↓** | ** **↓** | *** **↑** | n.s. | n.s. |
| Citric acid | * **↑** | n.s. | *** **↓** | *** **↓** | *** **↑** | *** **↑** |
| Formic acid | n.s. | * **↓** | n.s. | n.s. | *** **↑** | *** **↑** |
| Lactic acid | n.s. | * **↓** | * **↓** | * **↑** | *** **↑** | *** **↑** |
| 3-Hydroxybutyric acid | n.s. | *** **↓** | n.s. | *** **↑** | *** **↓** | * **↓** |
| Acetoacetic acid | n.s. | ** **↓** | *** **↑** | *** **↑** | *** **↓** | *** **↓** |
| Acetone | n.s. | n.s. | n.s. | ** **↑** | * **↓** | n.s. |
| Pyruvic acid | n.s. | ** **↓** | ** **↓** | *** **↑** | *** **↑** | *** **↑** |
